# A multimodal intervention for Alzheimer’s disease results in multifaceted systemic effects reflected in blood and ameliorates functional and cognitive outcomes

**DOI:** 10.1101/2022.09.27.22280385

**Authors:** Jared C. Roach, Lance Edens, Daria R. Markewych, Molly K. Rapozo, Junko Hara, Gustavo Glusman, Cory Funk, Jennifer Bramen, Priyanka Baloni, William R Shankle, Leroy Hood

## Abstract

**Introduction:** Comprehensive treatment of Alzheimer’s disease and related dementias (ADRD) requires not only pharmacologic treatment but also management of existing medical conditions and lifestyle modifications including diet, cognitive training, and exercise. The Coaching for Cognition in Alzheimer’s (COCOA) trial was a prospective randomized controlled trial (RCT) to test the hypothesis that a remotely coached multimodal lifestyle intervention would improve early-stage Alzheimer’s disease (AD). AD results from the interplay of multiple interacting dysfunctional biological systems. Specific causes of AD differ between individuals. Personalized, multimodal therapies are needed to best prevent and treat AD. COCOA collected psychometric, clinical, lifestyle, genomic, proteomic, metabolomic and microbiome data at multiple timepoints across two years for each participant. These data enable systems-biology analyses. We report analyses of the first COCOA data freeze. This analysis includes an evaluation of the effect of the intervention on outcome measures. It also includes systems analyses to identify molecular mediators that convey the effect of personalized multimodal lifestyle interventions on amelioration of cognitive trajectory.

**Methods:** A total of 55 participants with early-stage AD from Southern California were randomized into two parallel arms. Arm 1 (control; N=24) received standard of care. Arm 2 (intervention; N=31) also received telephonic personalized coaching for multiple lifestyle interventions including diet, exercise, and cognitive training. COCOA’s overarching aim was to gather dense molecular data from an AD cohort to improve understanding of pathophysiology and advance treatment. For the RCT, COCOA’s objective was to test the hypothesis that the Memory Performance Index (MPI) trajectory would be better in the intervention arm than in the control arm. The Functional Assessment Staging Test (FAST) was assessed for a secondary outcome. Assessments were blinded. The nature of the intervention precluded participant blinding.

**Results:** The intervention arm ameliorated 2.6 ± 0.8 MPI points (p = 0.0007; N = 48) compared to the control arm over the two-year intervention. Top-ranked candidate mediators included: albumin, propionylcarnitine, sphingomyelin, hexadecanedioate, acetylkynurenine, tiglylcarnitine, IL18R1, palmitoyl-sphingosine-phosphoethanolamine, acetyltryptophan, and IL17D. These individual molecules implicated inflammatory and nitrogen/tryptophan metabolism pathways. No important adverse events or side effects were observed.

**Conclusions:** Clinical trials should include frequent assessment of dense data to maximize knowledge gained. Such knowledge is useful not only in testing a primary hypothesis, but also in advancing basic biological and pathophysiological knowledge, understanding mechanisms explaining trial results, generating synergistic knowledge tangential to preconceived hypotheses, and refining interventions for clinical translation. Data from every trial should allow an intervention to be refined and then tested in future trials, driving iterative improvement. Multimodal lifestyle interventions are effective for ameliorating cognitive decline and may have an effect size larger than pharmacological interventions. Effects may be molecularly idiosyncratic; personalization of interventions is important. Dietary changes and exercise are likely to be beneficial components of multimodal interventions in many individuals. Remote coaching is an effective intervention for early stage ADRD. Remote interventions were effective during the COVID pandemic.

## 1. Introduction

Complex diseases require complex therapies.^1^ The COCOA trial was designed to test the hypothesis that multimodal interventions can slow progression, halt, or reverse — ameliorate — the course of Alzheimer’s disease and related disorders (ADRD). Intervention was delivered by telephonic coaching. This coaching leveraged knowledge from prior studies to choose interventions expected to ameliorate cognition, including diet, exercise, and cognitive training from BrainHQ (Posit Science Corporation). COCOA inclusion criteria encompassed individuals on the Alzheimer’s disease (AD) spectrum, centering on those with mild cognitive impairment (MCI). A shorter version of this manuscript intended as a quick read, skipping tangential discussion, technical details, and in-depth explanations is available on medRxiv.^2^ A discussion of the definition of AD and its relationship to COCOA inclusion criteria can be found in the Supplemental Introduction.

### COCOA epistemology

Our overall aims in designing the COCOA trial and other similar trials — such as PREVENTION (McEwen et al., 2021) — are to test and explore four broad overarching hypotheses that (1) there are multiple causes of AD, (2) there are multiple interventions to ameliorate AD, (3) different individuals with AD will have different mixes of causes, and (4) the best treatment for any given individual should be personalized to include multimodal interventions that treat the specific causes of their AD (**Figure 1**). Such interventions are best tailored to the individual’s personal circumstances (including genetic, environmental, and cultural influences). The best way to advance knowledge over all four of these broad hypotheses — and ultimately to provide specific clinically translatable knowledge — is a research program spanning many studies, many groups, and many years. However, most simple clinical trial designs would not be part of this research program, as they would provide only a narrow glimpse of the systems landscape. The COCOA trial designers acquiesced to tradition in that they pre-specified a simple hypothesis to be tested — that the intervention group will have a better MPI trajectory than the control group. This traditional aspect of the COCOA design enables some traditional approaches to epistemology. Notably, it allows us to avoid multiple-test corrections to the significance of our primary result. We will use this as a fundamental pillar of our claim that the COCOA intervention causes an amelioration of cognitive decline. We will bolster this claim with secondary outcomes (e.g., function) that are also improved. We will conclude that the COCOA intervention results in broad-based amelioration of AD trajectory.

**Figure 1.**
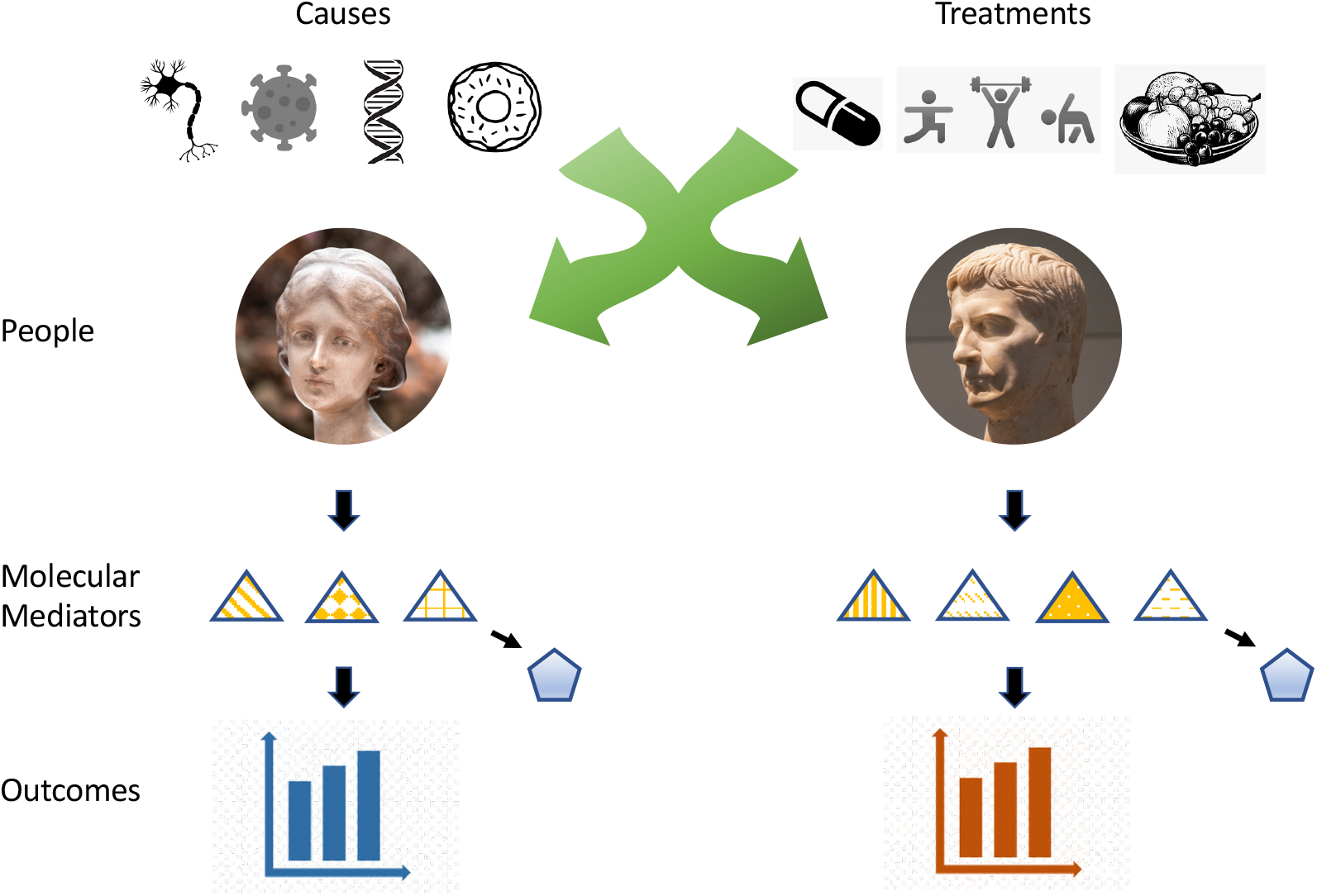
The multimodal nature of Alzheimer’s disease causes and treatments. Alzheimer’s disease (AD) has multiple causes and treatments. Each individual may have a different subset of these causes and receive a different subset of these treatments. A number of bioentities may mediate the effects of these treatments, including molecular mediators (triangles). Some of these same bioentities may also have mediated the cause of the disease. Outcomes for each individual depend on appropriately matching treatments to their individual pathophysiology. In this manuscript we focus on identifying mediators for treatments that ameliorate AD. In some cases, the effects of mediators may be measured by non-causal analytes that serve as biomarkers for these mediators (pentagons). Every individual is unique, but shared mechanisms — in aggregate across a population — unify the concept of AD.

However, this simple argument (lemma #1) does little by itself to advance any of our four overarching hypotheses. In order to advance these hypotheses, we will also show that (lemma #2) the COCOA intervention causes broad multisystem changes reflected in numerous blood analytes, (lemma #3) several of these analytes are correlated with cognitive improvement and are thus candidate mediators of the effects of the multimodal interventions, (lemma #4) there are mediators & mechanisms shared by multiple individuals that suggest some shared mechanisms may be broadly applicable to many or most AD patients, (lemma #5) there may be some mediators operating in only one or a few COCOA participants, (lemma #6) that the mechanisms identified — both aggregate and individual — are not randomly distributed across all systems, and (lemma #7) that several identified mechanisms have considerable prior evidence for mediating AD interventions.

Together these lemmas will bolster support for our four overarching hypotheses. They leverage the epistemological concepts of coherence — that the ensemble of the data fits together and are consistent with prior knowledge (Fedak et al., 2015). For COCOA, we need to modify the *coherence* epistemology criterion slightly. Our hypotheses assume multiple causes and therapies, so when we search for evidence of coherence, we do not expect a single unified coherent system, but several potentially distinct coherent systems. The overall syllogism is stronger than the sum of its parts. To a large extent, this syllogistic synergy stems from leveraging the coherence and plausibility elements of modern versions of Hill’s criteria (Fedak et al., 2015), specifically *biological plausibility*. This synergy is driven by the integration of knowledge from various evidence streams, particularly omics.

A recurring criticism of multimodal trials is that they should never be undertaken until all single components of the multimodal intervention have been previously tested individually in a portfolio of many unimodal intervention trials. We have addressed this concern with respect to COCOA and its sister trial (PREVENTION) elsewhere (e.g., Roach, Hara, Fridman, et al., 2022; Roach, Hodes, et al., 2022). Notably, such a research portfolio would take centuries, cost too much, and require more participants than are recruitable. In practice, embracing such a portfolio would mean no sufficiently complex multimodal intervention would ever be tested. Soon, it may be possible to test many thousands of hypotheses in a single clinical trial using modern high-throughput technologies and clever epistemology — COCOA can be considered a step in that direction. If indeed such a trial can be done on all proposed single interventions, then our argument against testing every intervention in isolation before including them in an omnibus trial would be weakened. However, some interventions have infinite variations, including diet and exercise (considering all doses, frequencies, and modalities): multimodal trials are inevitable and necessary even with improving technology and trial design. Therefore, COCOA is not specifically designed to be a parallel test of many individual components, but is also a synergistic test of a multimodal intervention — one that differs in particulars between participants due to personalization of the intervention. By collecting dense data, we plan to be able to improve recommendations for multimodal interventions by identifying molecular mediators (sometimes called ‘biomarkers’) that reflect mechanisms that identify and explain the effects of individual components of the interventions that are most beneficial in particular individuals.

Synergy may be important. If marooned on a desert island, one could insist on testing escape rafts built individually from available materials: driftwood, salvaged boards, coconuts, vines, and nails. All of these but nails might have some efficacy when used alone. But even using the best material in isolation might not result in a successful raft and the entire endeavor might be prematurely abandoned. The best raft is built from a multimodal combination of parts and includes nails — which in isolation do not even float. Systems epistemology is required to build the raft, leveraging both past knowledge of how rafts can be built (consistency) as well as on-island understanding of how the parts fit together in the current situation (coherency). Such epistemology also speeds up discovery. If getting off the island is urgent (perhaps there is no food or water — or even an impending volcanic eruption), then there may not be time to iterate through inefficient non-multimodal raft-building trials.

## 2. Methods

Methods for the COCOA trial were published as a separate manuscript (Roach, Hara, Fridman, et al., 2022). In brief, COCOA is a prospective randomized clinical trial (RCT) to test the hypothesis that coached multimodal interventions ameliorate cognitive decline. Participants were recruited from a high-volume memory clinic in Southern California. Demographics: 34% female; 92% White; average age 75 years. Main inclusion criteria: age 50 and older; Memory Performance Index (MPI; embic.us/application/clinical) of 65 or below. Main exclusion criterion: existing diagnosis of a non-AD neurodegenerative disorder. There were 55 participants randomized; attrition reduced the number to 35 who completed all 24 months of active participation in the trial. However, most individuals who did not complete 24 months participated long enough to be included in one or more of the analyses presented in this manuscript; N is indicated for each analysis in the corresponding sections (CONSORT flow diagram, **Supplemental Figure 1**). There were 8 females and 14 males in the control arm and 10 females and 21 males in the coaching arm. All regression and correlation p-values for the effects of the intervention and for the relationship of analyte changes to MPI score in **Supplemental Table 1** (and therefore all derivate tables in this manuscript) include sex as a variable. For this population, weight was 171 ± 35 lbs; BMI was 26 ± 4. At the time of trial design, power to detect an effect size of 1 MPI point of difference was estimated as 80% with N=36 participants. Assessments were blinded. The nature of the intervention precluded participant blinding. Participants were randomized into two arms using block randomization with variable-sized small blocks to achieve a 3:2 ratio of participants in the intervention arm to control arm. Informed consent was obtained from all participants. The trial protocol as approved by the Western Institutional Review Board (WIRB; protocol #20172152) is included as **Appendix 1**. A CONSORT checklist for clinical trials is included as **Appendix 2** (www.equator-network.org/reporting-guidelines/consort).

### Event Driven Procedural Modifications

In April 2019, Arivale, our primary provider of trial logistics, ceased operations. Most operations previously contracted through Arivale were then assumed in house. Enrollment ceased due to Arivale ceasing operations. Circa March 2020, the COVID pandemic began. COVID increased risk for in-person activities. Assessment interactions were rescheduled as necessary – in some cases creating a delay between planned assessments or causing a planned assessment to be skipped. The intervention — telephonic coaching — was exceptionally robust (almost presciently so) to COVID disruptions.

Unforeseen circumstances are likely in any long-term clinical trial. COCOA was well designed for these unforeseen circumstances. First, as mentioned above, COCOA’s interventions were driven by telephonic coaching, which was robust to the physical distancing (aka, “social distancing”) required by public health due to the pandemic. Second, COCOA’s design aimed at increasing diversity of data and interventions (Roach, Hara, Fridman, et al., 2022; Roach, Hodes, et al., 2022). Therefore, some increase in diversity of environment and experience is welcome, as long as there remains sufficient similarity between participants to produce common themes in systems analyses. By contrast, if a study *required* trial-participant and intervention homogeneity, then analyses of data from that study would have difficulty adjusting for universal environmental changes. In the case of COCOA, these changes happened unexpectedly as a result of the pandemic and procedural differences due to transitioning from a contracted provider to in-house provision of coaching. Since unexpected changes can be expected in any long trial, we endorse the COCOA trial design generally for such trials.

As illustrated in the Discussion of Roach, Hara, Fridman, et al. (2022), there are advantages to both diversity and homogeneity; a balance is needed between these extremes, and this balance may vary across different studies. In addition to our efforts to create diversity in our trial design, and embrace diversity resulting from unanticipated events, we also sought some homogeneity. In response to the cessation of Arivale, we hired coaching staff from Arivale in order to maintain provider continuity. Many of the COCOA assessments were obtained remotely, or could be, so there was little interruption or variation of these due to forced remote interactions. The MPI is a summary statistic of the MCI Screen (MCIS), which can be performed remotely.

One result of the event-driven changes was the reduction from a planned 200 participants (trial design) to 55 participants (trial completion). Of these 55, not all completed all planned assessments; some participants dropped out of the trial early and some participants skipped some assessments. Reasons for leaving the trial early included relocating out of state. No withdrawals were related to COVID, as far as we could ascertain. Most reasons for skipping assessments were because of physical-distancing needs and/or lack of health care staff to perform these assessments during intense COVID waves. This drop in the number of individuals and in longitudinal data points precluded calculating individual coefficients for participants in a linear mixed model (LMM); therefore we reverted to simple linear regression testing our primary endpoint. Use of a LMM was planned as specified in clinicaltrials.gov and in Roach, Hara, Fridman, et al. (2022). A change from a fully parameterized LMM to one with fixed slope for all individuals may or may not be considered a change from planned to implemented analysis algorithm depending on one’s viewpoint; but we highlight this change here as part of our “red team” effort to criticize our own work (Lakens, 2020).

### Recruitment

COCOA recruited, enrolled, and randomized 55 participants over 16 months, averaging over 3 enrollments per month (**Supplemental Figure 2**). The first participant was recruited in January 2018. The last participant was recruited in April 2019. Data collection from participants ended in Summer 2021, slightly longer than 24 months after the last participant started the trial; some assay collections were delayed due to the COVID pandemic. 42 participants completed at least two blood draws that were fully analyzed for both proteomic and metabolomic data; 43 completed at least two draws analyzed for the panel of clinical laboratory assays (aka, “clinical labs”) (**Supplemental Figure 3**). Enrollment was 92% self-declared White, reflecting the population at the enrollment site.

### Baseline Data

Baseline demographic characteristics are presented in **Table 1**. For this number of individuals in the two arms, the power to see a difference in a normally distributed baseline value between the two arms is 95% for a 20% difference in mean with 20% relative standard deviation. Statistical differences for the distribution of values between coaching and controls were compared by Fisher’s exact test for categorical values and two-tailed t-test for numerical values. We saw no evidence of imbalance between arms for any major demographic variable. Evenly allocating individuals exactly matching on every omics (or dense-data) variable between arms is impossible for any trial; COCOA did not attempt to balance arms for any particular variable. With a very large number of measurements, it is likely that at least some nominally significant p-values will be found in analyses, either at baseline or later analyses (Pusztai, 2007). Out of 1461 analytes, 144 had nominally significance (p < 0.05) for differences between baseline means; about 73 would have been expected by chance. Because of correlations between variables, we did not consider the difference between 144 and 73 to be exceptional. None of these 144 analytes had significance after Bonferroni correction. Thus, although randomization was effective, not all baseline distributions of specific analytes were equivalent between the two arms. A relatively slight variation in a demographic variable, such as all four non-White participants in the coaching arm, could conceivably result in a significant bias in multiple molecular baseline variables, particularly if baseline diet were correlated with a particular demographic variable. Overconfidence in results (e.g., Type I error) due to regression toward the mean may be more likely in analytes with baseline differences and molecular systems incorporating them. Such baseline differences are likely in all trials but may not be recognized if dense baseline omics data is not collected. It is also formally possible that slight variations could have been introduced between the two arms in the days (in some cases weeks) after randomization but before the baseline blood draw, as participants were aware of their assignment. However, coaching did not occur prior to this blood draw. All blood draws were morning fasting draws; they should not have a circadian or length-of-fast bias, but we cannot rule out a subtle effect, particularly on metabolites sensitive to length of fast (e.g., Solianik et al., 2022).

## 3. Results and Intermediate Logical Steps (Lemmas)

We present our result in seven subsections, each corresponding to a lemma proposed in the Epistemology section of the Introduction. In brief, lemma #1 is our claim that the COCOA intervention results in cognitive and functional improvement. The other lemmas are: (lemma #2) the COCOA intervention causes broad multisystem changes reflected in numerous blood analytes, (lemma #3) several of these analytes are correlated with cognitive improvement and are thus candidate mediators of the effects of the multimodal interventions, (lemma #4) there are mediators & mechanisms shared by multiple individuals that suggest some shared mechanisms may be broadly applicable to many or most AD patients, (lemma #5) there may be some mediators operating in only one or a few COCOA participants, (lemma #6) the mechanisms identified — both aggregate and individual — are not randomly distributed across all systems, and (lemma #7) several identified mechanisms have considerable prior evidence for mediating AD interventions.

### 3.A. Lemma 1: The COCOA intervention causes an amelioration of cognitive and functional trajectories

One of two major elements of the design of the COCOA Study was to craft an RCT to test the hypotheses that the COCOA intervention causes an amelioration of cognitive (primary hypothesis) and functional (secondary hypothesis) trajectories. We leverage the RCT design and results to demonstrate that these two hypotheses were correct.

#### Cognition (MPI) is significantly ameliorated in the intervention arm

Linear regression showed an effect size of a 2.6 ± 0.8 MPI point benefit (p = 0.0007; N= 19 control and 29 intervention, 48 total) to the intervention (coached) arm compared to the control (standard of care) arm over the two-year course of the intervention (**Supplemental Figure 4**). We report one-sided significance; we pre-declared our hypothesis that the intervention arm would have a better outcome than the control arm. We performed a similar analysis with the MoCA. There is a trend towards ameliorated MoCA in the intervention arm, but it is not significant (**Supplemental Figure 6**). We expected less power for MoCA, as we measure MoCA less frequently and it has less precision than the MPI, so this lack of MoCA significance is not surprising (Roach, Hara, Edens, et al., 2022). The MPI benefit was most evidenced as a fixed effect, rather than as an interactive term with time, suggesting that the trajectory of benefit was nonlinear. We fit a spline curve to the data to illustrate this non-linearity (**Figure 2**). This spline shows that most of the cognitive benefit occurs in the first year of intervention – perhaps during the first several months – and that this benefit is maintained as a fixed (rather than increasing or decreasing) difference from the cognitive trajectory of the control arm. This benefit remains for the duration of COCOA, and perhaps beyond — at least two years after the commencement of intervention. Indeed, even though there is, over a two-year average, a negative trend to the cognitive trajectory of the COCOA intervention arm, this trajectory does not significantly dip below the baseline. Therefore, we cannot reject the claim that the participants in the intervention arm in aggregate had no cognitive decline, although we are not highly powered to test that claim. COCOA was powered to test the null hypothesis that the intervention arm was not better than the control arm — and was able to significantly reject that null hypothesis. Although the effect is nonlinear, and therefore not easily apportioned over time as rate (or difference between rates) of improvement, it is important to convey a sense of the magnitude of the effect, so that it can be compared to effect sizes of other interventions for AD. The average change in MPI score between enrollment and end of the trial (withdrawal, death, or completion), for those individuals with at least two assessments, was −4.6 ± 3.2 (N=29) in the control arm and − 1.7 ± 3.1 (N=19) in the intervention arm, which is very approximately 60% less cognitive loss. This effect size eclipses known effect sizes from pharmaceutical interventions (the best typically 10-20%) and exceeds most reported effect sizes from non-pharmaceutical interventions (the best typically 46-59%) (Hendrix, Nicodemus-Johnson, Hogan, et al., 2022). Making such comparisons is difficult because of differences in outcome measures and other aspects of trials, so we make this claim very cautiously. Indeed, our reported effect size, as well as the reported effect sizes we cite for comparison, may be inflated (Ioannidis, 2008). Furthermore, if trajectories are non-linear and/or have oppositely signed slopes for different arms, as portrayed in **Figure 2**, it is not clear that discussion of “effect size” is a meaningful way to present results — or at least it becomes important to use tailored definitions of effect sizes for each trial. Tailored effect sizes can be based on time delay (Raket, 2022) or area between trajectories, and thus it is hard to compare effect sizes among and between all trials. For example, had we planned to end the trial at a different timepoint than 24 months, we could have observed a very different effect size and/or been forced to use a different definition of effect size.

**Figure 2.**
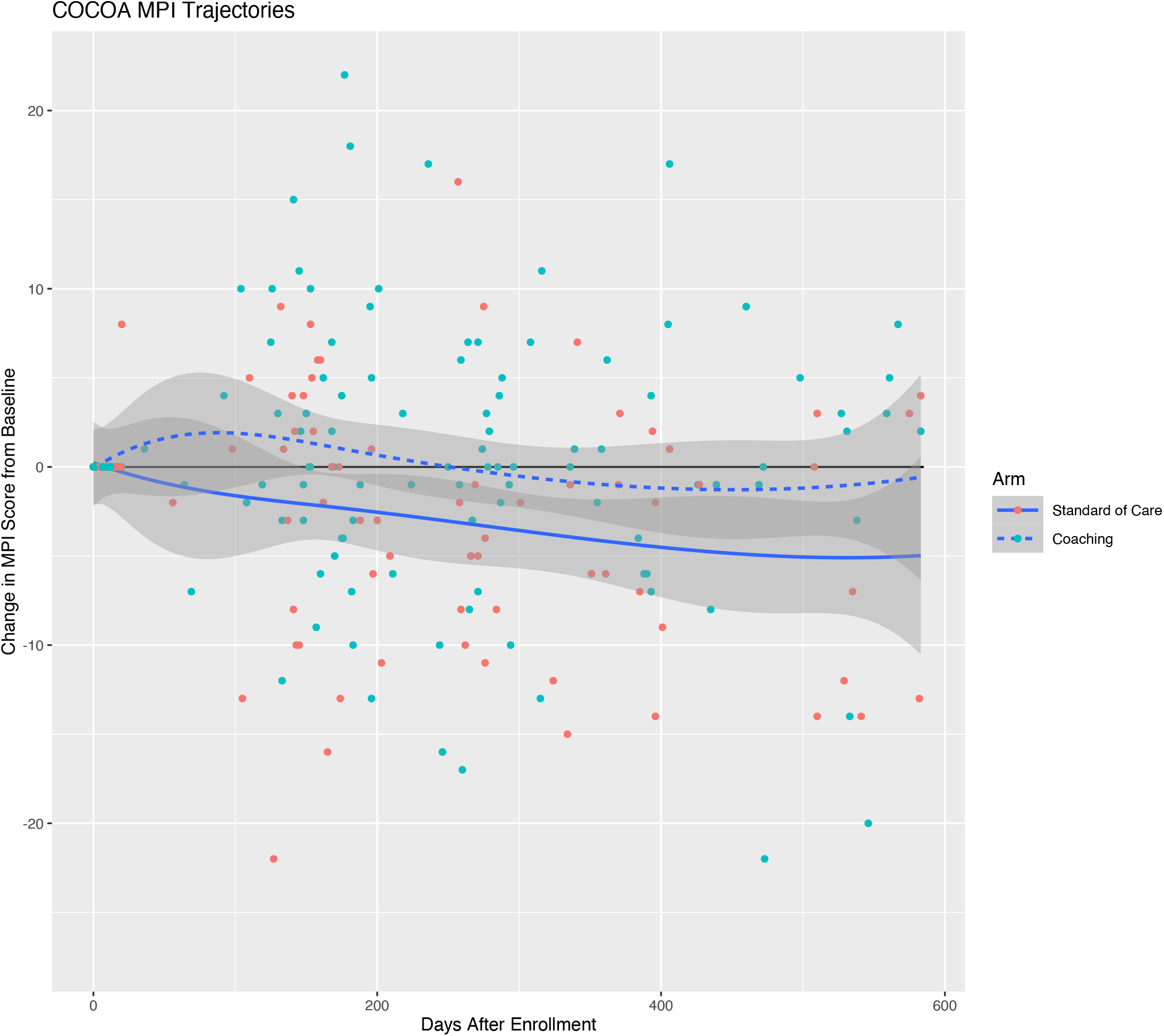
Aggregate MPI trajectory of COCOA Participants. The COCOA intervention ameliorates aggregate cognitive trajectory. Each MPI measurement from every participant is shown. Cognitive measurements vary considerably between individuals and over time. Change from baseline is graphed. The primary outcome measure is computed with linear regression and graphed in **Supplemental Figure 4**. However, as the aggregate cognitive trajectories of each arm are most likely non-linear, this figure (illustrated with spline trajectories) may more accurately convey the nature of the trajectories than a linear fit. The baseline is shown as a dark black line and serves as a reference for no change over time. The intervention arm diverges markedly from the control arm during the first several months of the trial, and then maintains a fairly constant improvement over the remaining months. Both trajectories eventually drop below baseline, but with the intervention arm losing relatively little cognition from baseline. Individual trajectories are graphed in **Supplemental Figure 5**. 90% confidence intervals are shaded.

#### Effects of sex, age, and education

Sex, age, and education influence cognitive assays in many contexts. We performed linear regression including terms for sex, age, and education. Of these, only sex is an independent predictor of MPI trajectory in COCOA (two-sided p-value = 0.0064), with female sex having an aggregate beneficial effect (2.2 MPI point benefit). COCOA is underpowered to observe effects of age (p = 0.29) and education (p = 0.29), if any, on MPI trajectory. The interactive term between sex and arm is not significant, indicating that the effects of the COCOA intervention and sex are independent. There are no large differences in the distributions of sex, age, or education between arms (**Table 1 & Supplemental Table 1**).

#### Function (FAST) is significantly ameliorated in the intervention arm

Linear regression showed a benefit to FAST (one-sided p = 0.030), a secondary outcome measure (**Supplemental Figure 7)**. Individuals with at least two assessments in the control arm deteriorated (i.e., increased) 0.53 ± 1.2 (N=15) FAST score points over the course of the trial and individuals in the intervention arm deteriorated only 0.33 ± 1.1 (N=27) FAST score points, very approximately a 40% reduction. Therefore, both a measure of cognition (MPI) and a measure of function (FAST) ameliorated due to the COCOA intervention. This suggests that the COCOA intervention has broad multifaceted benefits across multiple systems — that the COCOA intervention is beneficial for overall brain health and for patient-oriented outcomes relevant to dementia.

The robustness of the FAST result presented in **Supplemental Figure 7** is mediocre. For example, if the three participants with the largest absolute changes in FAST score are removed from the analysis, the p-value for the improvement in cases versus controls would no longer be nominally significant. Although the significance is not robust, the trend is: one has to remove the 10 participants (out of 42 with at least two FAST measurements) with the largest absolute changes to eliminate the trend toward aggregate improvement. This robustness analysis implies that the largest benefits from the intervention may be accrued by a relatively few individuals, that there may be a small benefit enjoyed by many participants, and that at least some individuals in the intervention group may receive no benefit. It is also possible that our FAST improvement is a chimera resulting from a type I error.

FAST is a low-precision instrument with very large patient oriented (and undoubtedly underlying pathological) differences between consecutive categorizations. For example, FAST stage 2 is “subjective functional deficit”; stage 3 is “objective functional deficit interferes with a person’s most complex tasks”; and stage 4 is “instrumental activities of daily living (IADLs) become affected, such as bill paying, cooking, cleaning, traveling”. In untreated AD, it typically takes several years to degenerate from FAST stage 3 to 4, and two years to degenerate from FAST stage 4 to 5 (Reisberg, 1986). It is rare for FAST to improve. It is therefore particularly interesting to compare the number of observations of FAST improvement between cases and controls. We considered all pairs of consecutive FAST assessments of the same individual. In control participants, 1 of 32 (3.1%) such intervals showed improvement; in intervention participants, 6 of 65 intervals (8.6%) showed improvement (**Supplemental Figure 8**). This difference is not significant by Fisher’s exact test, but the magnitude of the difference suggests that the intervention may occasionally reverse the process of disease (or at least reverse the decline of patient-oriented outcomes) in some individuals.

Considering just the FAST and MPI p-values, there are two extreme approaches to combining significance. Both of these are fallacious (Theiler, 2004), but serve to define extremes. The first approach is to consider these two p-values as testing two distinct hypotheses (Hypothesis A: MPI trajectory is identical; Hypothesis B: FAST trajectory is identical) and divide each significance by two to account for “multiple testing”. Since we pre-declared a single primary outcome measure, this would be unconventional. The second approach is to consider the p-values to result from completely independent tests of the same hypothesis (that dementia trajectory is identical) and multiply the p-values. Leveraging Bayesian epistemology, such a multiplication might include a prior probability – but it should also include factors for all evidence, both advancing and retarding the hypothesis, which is not an endeavor that is currently feasible. It would also have been possible to use a prespecified bivariate hypothesis to combine p-values non-multiplicatively, which is not fallacious, but is unusual (Theiler, 2004). Depending on perspective, leaning towards either of these extremes could increase or decrease confidence in an overall beneficial effect of the intervention. However, as described below in our molecular results, we prefer to integrate all data (including the >1,000 molecular data points) before firming an opinion on the significance of an overall effect of the intervention.

We consider the above evidence to establish lemma #1. We bolster this evidence further with a comparison to cognitive trajectories observed in a large cohort study: ADNI.

#### Comparison to ADNI trajectories

Our primary outcome linear model for COCOA estimates a −0.2 MPI point decline per year in cases and a −2.8 MPI point decline per year in controls. In the Alzheimer’s Disease Neuroimaging Initiative Database (ADNI) cohort, in aggregate, MoCA scores decline about −2.2 points per year in AD participants (**Supplemental Figure 9**). In COCOA data, the MoCA score and MPI score are correlated: a one-point change in MoCA corresponds to a 0.31 point change in MPI (data not shown). This coefficient fits intuition, since the maximum MoCA score is 30 and the maximum MPI score is 100. Using this coefficient to convert COCOA’s MPI predictions to MoCA equivalence, our primary outcome linear model estimates a −0.1 MoCA point decline per year in cases and a −0.9 MoCA point decline per year in controls. COCOA controls receive standard of care. ADNI participants presumably receive standard of care, although ADNI is not an interventional study. Therefore, it is expected that both groups have similar aggregate decline. This difference (COCOA controls’ −0.9 and ADNI’s −2.2 MoCA points) is arguably indeed similar, but is noticeable. The difference between these two values is within our comparison’s margin of error. It is also likely that on average, COCOA controls do better than ADNI participants. Possible explanations: COCOA participants receive care at specialty memory clinics and/or benefit from the specific environment and demographics of Orange County (e.g., affluence), and ADNI participants receive care from diverse providers, including those not specializing in memory care, and reside in diverse locales.

External datasets like ADNI are useful for validation and for strengthening the epistemological pillar focused on consistency — the comparisons establish that COCOA results are consistent with the results of other research (Fedak et al., 2015). The difference in the regression coefficient for cognitive decline between COCOA controls and ADNI participants highlights one fallacy of categorizing individuals in an external dataset as “controls”. Unless controls are drawn from the same population as cases, an overt or hidden confounder may distort comparisons. In theory, an accurate systems model could completely account for confounders, but (1) we do not yet have such a model, and (2) if one had such a model it is not clear why more research requiring such external controls would be needed, except if it was fully accurate at modeling the system in control conditions but not able to model the system in intervention conditions, so use cases would be limited.

#### Minimal Clinically Important Difference

The minimal important difference (MID) or minimal clinically important difference (MCID) is the smallest change in a treatment outcome that an individual patient would identify as important or would indicate a change in the patient’s management. This inherently subjective metric can be estimated objectively; one estimate of the MCID for MoCA is 1.22 (Wu et al., 2019). Over the two-year course of COCOA, we observe about a 1.6 point difference in MoCA between arms, which would be clinically significant by this metric.

### 3.B. Lemma 2: The COCOA intervention causes systemic molecular changes in the blood

We expect that the COCOA intervention causes many effects in many organs and tissues, and that these are reflected in the blood. Indeed, it could be argued that the prior knowledge accumulated over more than a century (and resulting prior probability) of the influence of altering multiple lifestyles, including diet and exercise, is sufficient to make this claim without any further evidence. Such lifestyle alterations will certainly cause many changes in the blood. Therefore, we could, in principle, skip proof of this lemma, given that it is so obviously true. However, it might be argued that our intervention (coaching) is ineffective at producing lifestyle changes, and that therefore no blood molecular changes are expected. Indeed, this argument has been made (Daly, 2022). Therefore, it is worth demonstrating with actual COCOA data. The resulting analysis also reveals which analytes and subsystems are altered.

In this manuscript, we analyze a subset of data produced from the COCOA trial. This subset consists of three classes of blood omics data: plasma proteomics, plasma metabolomics, and clinical labs. Collectively, we call all the measured molecules and metrics from these assays “analytes”. Proteomics were contracted to Olink Bioscience (Uppsala, Sweden) and consisted of five chip-based targeted proteomic panels: NEU, NEX (Reagent: Olink Target 96 Neurology Panels; Source: www.olink.com/products-services/target/neurology-panel; Identifiers: NEU & NEX), INF (Reagent: Olink Target 96 Inflammation Panels; Source: www.olink.com/products-services/target/inflammation; Identifier: INF), CVD2, and CVD3 (Reagent: Olink Target 96 Cardiovascular Panels; Source: https://www.olink.com/products-services/target/cardiometabolic-panel; Identifiers: CVD2 & CVD3). These five Olink panels encompass 456 distinct protein assays targeting 443 distinct proteins chosen by Olink for relevance to neurobiology, cardiovascular disease, and inflammation. Olink protein concentrations are reported in Normalized Protein eXpression units (NPX, https://www.olink.com/faq/what-is-npx); these are batch-corrected in a pre-processing phase of our workflow. Metabolomics were contracted to Metabolon (Reagent: LC-MS global metabolomics platform; Source: www.metabolon.com/solutions/global-metabolomics; Identifier: Meta GA). Metabolon’s untargeted mass spectrometry typically detects >1000 metabolites in an individual plasma sample. Metabolites found in few samples were excluded (i.e., a metabolite must be seen longitudinally — including at least the first two timepoints — in 25% of individuals with data); after these exclusions, 952 metabolites were left to be included in our global analyses. Metabolon metabolite concentrations are reported in units of relative abundance (Busch, 2001); these are batch-corrected in a pre-processing phase of our workflow. Clinical labs were contracted to LabCorp; 53 clinical labs are included in this analysis; excluded labs either had little variability (e.g., almost always zero) or were highly correlated with an included lab — typically the calculated values like glomerular filtration rate (GFR). One clinical lab — urea (aka, blood urea nitrogen or BUN) — is also assayed by our Metabolon analysis. Clinical labs are reported with units standard for each in the United States (www.labcorp.com/test-menu). Detailed methods are described in Roach, Hara, Fridman, et al. (2022). Out of 55 randomized participants, 48 had at least one set of clinical labs, and 42 had at least two sets. Unless otherwise specified, 1461 analytes are included in our global analyses, encompassing 456 protein assays, 952 metabolites, and 53 clinical labs: 1447 of these are distinct.

In this manuscript, we focus on the initial molecular physiological trajectory of participants. There are three major reasons for this focus. First, we are reporting on the first COCOA data freeze, which includes these data. Second, our analysis of the aggregate cognitive trajectory (**Figure 2**) suggests that the response to intervention is non-linear, with the greatest divergence between the case and control trajectory happening during the first six months of the intervention. This corresponds to the length of time between the baseline and first follow-up omics blood draw. Many other AD interventions — such as donepezil (Burns et al., 2007) — also show non-linear responses, with the major effect occurring in the first six months, so this nonlinearity was expected at the time we designed the trial — but not so strongly that we would have felt comfortable proposing a non-linear test of our primary hypothesis. We do not rule out the possibility of continued omic difference between cases and controls past six months. Indeed, although the differences in aggregate cognitive outcome trajectories remain parallel rather than continuing to diverge after six months, this difference may be maintained through continued dynamic molecular physiologic changes. However, we defer that analysis for future research. Third, some individuals left the trial and some blood draws were skipped. Therefore, analytical power is greatest for the baseline to first timepoint compared to all other pairs of timepoints. Thus, omics values for the major analyses presented in this manuscript are the difference between baseline and the first timepoint for each individual (timepoint 1 value minus baseline value). This choice has the benefit of controlling for many potential confounders. Any factor with an invariant effect on an individual over the period of analysis will be eliminated. However, not all potential confounders will have invariant effects. Age affects the blood proteome (Earls et al., 2019) but only gradually so is unlikely to be a confounder over our six-month interval. Sex is invariant in our cohort, but particularly since it influences MPI trajectory (see above), the *influence* of sex might not be invariant (e.g., on average, sex may impact responsiveness to coaching or molecular changes in response to exercise). For aggregate analyses, randomization between arms mitigates the effects of confounders.

There are multiple approaches for evaluating the effect of an intervention on a molecular system (or a set of such systems). One is to predefine a set of subsystems (e.g., Gene Ontology), and then to use omics data to determine if there is a significant influence of an intervention on that subsystem. Gene-set enrichment analysis is an example of such an approach. Another approach is to allow the data to define the systems (and subsystems), as well as to illuminate any significant changes to those systems in response to intervention. We use both approaches to develop our first argument that the COCOA intervention causes systemic molecular changes in the blood. We start with the second approach: learning clusters from our own COCOA data.

First, we computed correlations between the 1461 analytes with sufficient data. These correlations were computed by comparing values of the two analytes between individuals, where the value of analyte for an individual is the level of the analyte at their first follow-up assay minus their baseline value for that analyte. We converted these correlations to distances as the absolute value of the complement of the correlation (abs(1-cor)), and dimensionally reduced this distance matrix. We converted correlations to absolute values because we are interested in shared information between analytes.

We used two dimensionality reduction approaches. The first, principal coordinates analysis (PCoA or “classical”) multidimensional scaling (MDS), emphasizes the effects of the multitude of all correlations of each analyte, and therefore is dominated by many small effects. The second, thresholded force-directed network layout, emphasizes only the strongest correlations, creating a more interpretable network.

Frequently in systems biology, clustering analyses are based on correlations of analytes across many conditions (for example, culturing genetically identical bacteria in many different media), and therefore are likely to produce clusters and networks that reflect biological function/mechanism/pathway. In the case of COCOA, we are computing correlations across many individuals participating in COCOA. Although clustering likely represents function, it may not. At least three factors could potentially contribute to a correlation between two analytes: (1) they may be functionally related, (2) they may be caused by the same factor, and (3) they may be the result of systematic error such as a batch effect. By way of illustration, in hot weather, one may observe an ice cube melt, a dog pant, and a dog jump into a lake. All of these events are correlated. The panting and the jumping are caused by the same factor (heat) and are functionally related — they serve to cool the dog. The melting and the panting are caused by the same factor (heat) but are not functionally related. We expect one factor driving strong correlations between analytes to be the COCOA intervention because our trial intervention was designed to perturb molecular systems, and we know from our analysis presented above in lemma 2 that many analytes are affected by arm assignment. Also, we have carefully controlled for batch effects; individuals and samples are randomly distributed between technical batches, and pooled controls are used for all normalizations. Despite these controls, there are other systematic influences that cause more clustering than would be expected at random. In particular — similar analytes tend to cluster together (i.e., proteins with proteins and metabolites with metabolites). And although it might be the biggest factor, the COCOA intervention (arm assignment) might not be the only factor driving clustering. Sex or an environmental effect orthogonal to the coached intervention might be a factor — so we would not expect all clusters (“subsystems”) in our correlation analyses to be explained/altered/affected by COCOA arm assignment. We have yet to fully quantify some of these effects (e.g., by building a simulation model), so currently judgment of the significance of clustering remains somewhat subjective.

In addition to computing all-by-all correlations, we computed two statistics for each analyte. One statistic estimates the analyte’s value as a biomarker for the intervention, and the other statistic estimates the analyte’s value as a biomarker for the outcome. These two “intervention value” and “outcome value” statistics were computed as the significance of the slope of the regression between the analyte value and: (statistic 1) arm assignment or, (statistic 2) the overall MPI slope. Both statistics include an adjustment for sex (i.e., the regressions include sex as an explanatory variable). For each analyte, the value for each participant is the level of the analyte at their first follow-up assay minus their baseline value for that analyte. Overall MPI slope is computed as their final MPI score minus their first MPI score, and then divided by the number of days between those two MPI assessments. Overall slope is a better indicator of clinically relevant outcome than slope between the first two timepoints. Each regression analysis for each analyte was performed separately. Two significances were computed for each analyte from two separate regressions: one including all data (“full analysis”), and one excluding outliers (“robust analysis”). Grubbs’ test was used to identify analytes with outliers. If Grubbs’ test significance was < 0.05, the most extreme value (i.e., participant) was excluded from that analyte from the “robust” analysis. We focus on the intervention statistic in lemma 2, as our purpose in this lemma is to show that the intervention has an effect on blood omics.

To provide evidence for the causal effect of the COCOA intervention on the blood, **Figure 3** is an MDS representation of all analytes. The significance of association between the case and control values for that analyte is represented by the size of the icon. The clustering of the significant analytes is astronomically significant by inspection. This clustering is unlikely to be due to batch effects, because: (1) the biggest cluster enriched in significant analytes includes all three categories of analytes, each of which was assayed by a different methodology, (2) we used rigorous batch correction techniques for our Olink and Metabolon assays, beyond what is recommended by the vendors, by including multiple control samples in all batches, and (3) batches were not based on the individual’s timepoint or arm, but rather on real-time accumulation of samples which spanned many timepoints for different individuals, as they enrolled at different real times. Therefore, the clustering must be due to the intervention. Therefore, the intervention causes manifold changes in analytes in the blood.

**Figure 3.**
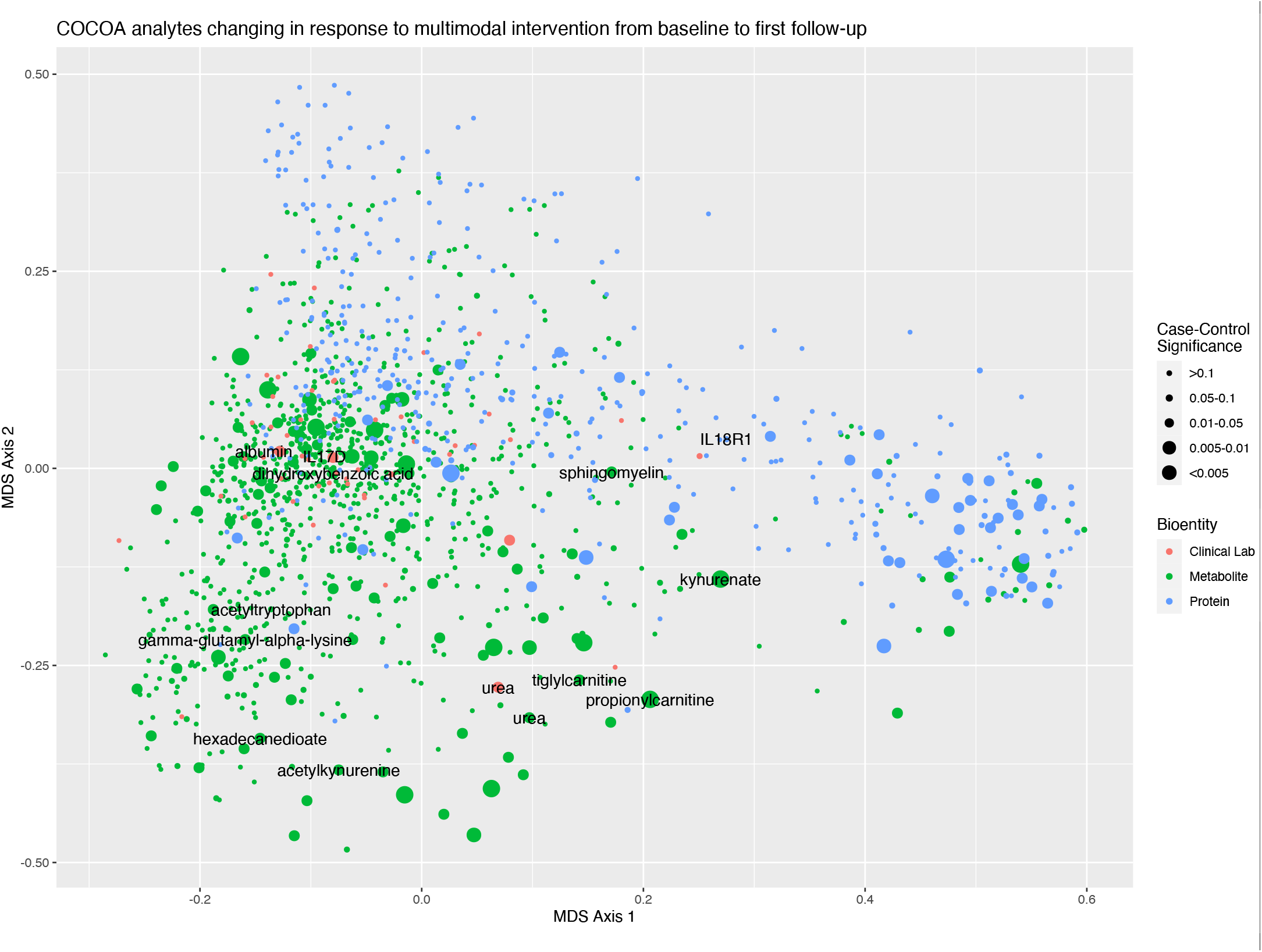
The COCOA intervention causes systemic changes in multiple molecular subsystems reflected in the blood. This visualization results from dimensionality reduction of the distances between all COCOA analytes as determined by absolute correlation distances. Significant nodes cluster, which implies that the effects of the COCOA intervention are not randomly spread across biological systems and may be targeted to systems specifically impacted by the intervention; random distribution would be expected if results were pure noise and no signal. The principal coordinates analysis (PCoA) multidimensional scaling (MDS) algorithm was used for dimensionality reduction. This algorithm incorporates all correlations; the resulting layout is dominated by the many effects of multiple small correlations, which may in aggregate contribute more to the placement of a node than the strongest correlations to that node. Larger nodes indicate significance of the effect of the COCOA intervention. Labels are added to nodes that are discussed in the main text of the manuscript. Urea appears twice, once as a clinical lab measurement, and once as a metabolite measurement.

We also performed an analysis focused solely on metabolite analytes (**Figure 4**). This approach leverages pre-compiled lists and permitted us to use an off-the-shelf metabolite set-enrichment bioinformatics tool. Focusing on a single data type also reduces certain statistical concerns associated with combining datasets with very different units and properties of measurement distributions. The systems were represented by these analytes included nitrogen metabolism, bile acid, and xenobiotic pathways.

**Figure 4.**
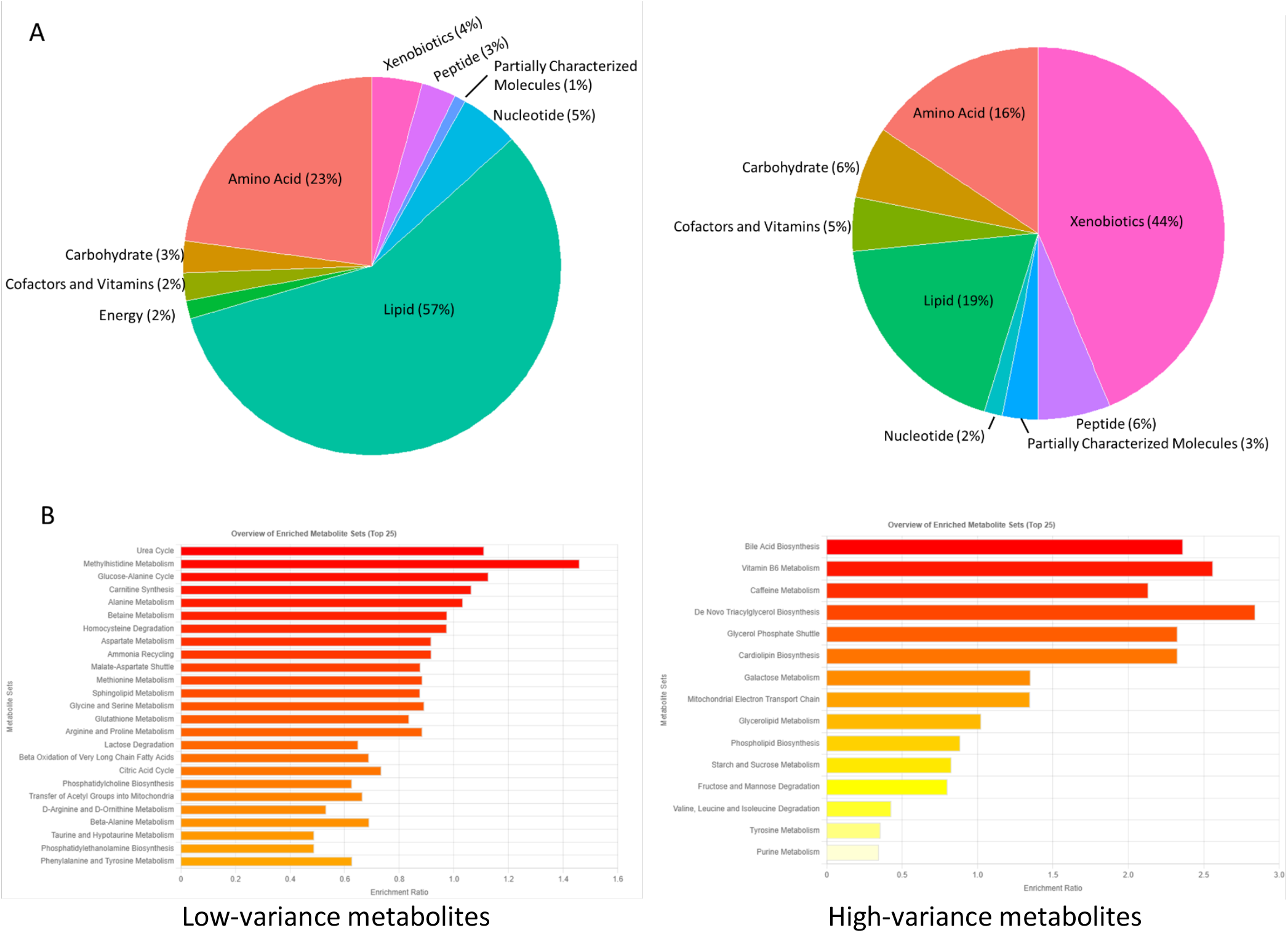
Bile acid, xenobiotic, and nitrogen metabolism pathways are likely altered by the COCOA intervention. The top pathways implicated by alterations in metabolites in response to the COCOA intervention are all consistent with prior knowledge that these pathways are impacted by diet and consumption of xenobiotics. Some of these pathways are also known to be affected by exercise and other components of the multimodal intervention. In order to leverage an existing workflow for pathway enrichment (Metaboanalyst, www.metaboanalyst.ca; Pang et al., 2021), for this figure only, we limited our analysis to the metabolite subset of all COCOA analytes. We divided metabolites into two groups, based on overall variance of all measurements of that analyte. High-variance analytes might have non-monotonic and non-linear trajectories; sampling such trajectories a few times a year could abet overconfidence. Because power to detect the effect of the intervention on a pathway depends on how well the components of that pathway can be detected by metabolite mass spectrometry and COCOA sampling frequency, failure to highly rank a pathway is not strong evidence for the absence of effects of the intervention on that pathway. (A) Pie charts indicate the category abundance of metabolites in each classification. (B) Pathway enrichment. (Left panel) Nitrogen metabolism pathways are enriched by low-variance metabolites; (Right panel) Bile acid and xenobiotic pathways are enriched by high-variance metabolites.

We are using a multimodal intervention. Although the desire is for this intervention to be as personalized as possible, individualization of precision medicine approaches is still developing. Therefore, many aspects of our interventions — or perhaps entire categories of intervention — will have no effect on cognition in any given — or perhaps every — individual. Furthermore, even if we know what node or subsystem we want to target (e.g., blood pressure), few interventions specifically target that element of the system. For example, exercise may reduce blood pressure, but may also exacerbate arthritis. If we were to describe multimodal interventions hyperbolically, we might write that we were “throwing everything but the kitchen sink” at AD. Therefore, we expect many of the molecular effects of our current multimodal intervention to have no consequential/causal downstream effect on dementia/cognition/function. Our demonstration in this section — that the COCOA intervention has many molecular effects on blood analytes — does not demonstrate that all — or even any — of them influence dementia/cognition/function. Therefore, although we have strong — if not definitive — evidence of lemma 2, in order to advance our overall hypotheses, we need to present arguments for the remaining lemmas, as described in the following paragraphs.

### 3.C. Lemma 3: Molecular changes in the blood of COCOA participants are correlated with cognitive improvement

For this lemma, we do not consider participant arm; that is, we ignore the case or control “label”. We treat the COCOA trial as a cohort study. Individuals in the study differ in lifestyle for many reasons; they differ genetically and environmentally. Therefore, analyses for this lemma may identify associations with analytes that are not targeted by the COCOA intervention, as well as those that are.

The analytes most correlated with overall MPI slope are presented in **Table 3**. Xenobiotics, such as aspirin and food components (in some cases, “supplements”), are common among the most significant analytes. The salicylate and N-acetylalliin (a xenobiotic metabolite that can be derived from garlic) are among the top five analytes for both the “full” and “robust” analyses. Aspirin dose is a known predictor of cognitive trajectory in AD, at least in some contexts (Wattmo et al., 2011). The prominent presence of xenobiotics in these lists suggests that interventions other than the COCOA intervention may play a large role in cognitive amelioration. The effect of these xenobiotics appears to be independent of the effect of the multimodal lifestyle intervention. Therefore, the best interventions for dementia may include combinations of both lifestyle and pharmacological/xenobiotic interventions. Consumption of particular pharmaceutical or dietary chemicals may be correlated with cognitive trajectory but may or may not be causal. For example, a non-exclusive explanation for the significance of these xenobiotics is that the presence of xenobiotics in the blood is a biomarker of attention and compliance to health advice — these xenobiotics may *correlate* with cognitive improvement but not *cause* cognitive improvement. We do not revisit discussion of these xenobiotics in this manuscript, but they augur value of a future manuscript to focus on consideration of the COCOA data from the perspective of a cohort analysis, rather than of an RCT.

Some true positives, if they exist, may be easier to identify than others. For example, subtle changes in the baseline value of highly periodic (e.g., circadian) analyte could be hard to discern without very dense data. In the present analysis we are not adjusting p-values or expected discovery rates for individual analytes. In theory, power for each analyte could be separately estimated based on expectations of the noise contributing to measurements of that particular analyte (ranging from effect of circadian rhythm to technical aspects of the molecular assay). In practice, such power analysis is difficult to perform with enough confidence to enhance our overall argument.

Including an adjustment for sex had little effect on the ranking of analytes for analyses of this lemma. The average difference in rank was 5.8 for the “full” analysis among analytes in the top 50 — either including sex as an explanatory variable or not. A completely random reordering of lists of length 1461 would result in an average distance between ranks of 487. We conclude that if there are true associations with cognitive trajectory among those analytes we identify as most significant in aggregate, these particular associations are likely to be present in both sexes.

### 3.D. Lemma 4: Some potential mediators & mechanisms may be shared by multiple individuals

A potential mediator should (1) be influenced by the COCOA intervention, (2) influence cognition, and (3) the intervention effect should be in the same direction as the cognition effect. Therefore, analytes identified as top candidates in *both* lemma 2 and lemma 3 are potential mediators. Although our overarching hypothesis is that there are multiple mechanisms that may contribute personalizable therapies for AD, and that these may not be shared between particular individuals, we are confident there will be some that are shared between many individuals diagnosed with AD. If that were not the case, AD would no longer merit distinction as a named disease entity. Therefore, we aimed to detect shared or common molecular mediators of the COCOA intervention. To this end, we compared the top-ranked “robust” rankings of analytes that ranked highly in both (1) significance of response to the COCOA intervention (listed in **Table 2**, from lemma 2), and (2) significance of association with MPI slope (listed in **Table 3**, from lemma 3). These include both “full”-data ranking and “robust” rankings. The intersection between these lists is shown in **Table 4**. Three of the top seven analytes (kynurenate, N-acetylkynurenine, N-acetyltryptophan) are associated with tryptophan metabolism. This coherence suggests that certain subsystems (in addition to individual analytes) might mediate the intervention.

There are four analytes that are nominally significant in both Table 2 and Table 3 (full analysis): propionylcarnitine, kynurenate, gamma-glutamyl-alpha-lysine, and albumin. This overlap of 4 is about the same as would be expected solely due to the simplest prediction from type I error (0.05 × 0.05 × 1461 = 3.6). Even though our p-values are approximate — and we likely lack power to detect changes in many of the analytes due to analyte idiosyncrasies (e.g. sparse consumption of certain xenobiotics), lack of much greater intersection than might be expected by chance weakens our argument that there is true signal in this list; our argument must rely on coherence and consistency considerations, which we describe in lemmas 6 & 7, below.

### 3.E. Lemma 5: Some potential mediators may be operating in only one or a few COCOA participants

It is possible that some individuals may respond particularly well to a specific component of the COCOA intervention and/or via a specific molecular mediator. COCOA is underpowered to convincingly detect such “personal mediators” solely through univariate analyses. Nevertheless, an ensemble of candidate personal mediators might show intra-ensemble coherence and/or coherence with the more global mediators identified in the previous subsection. To this end, we compared the top-ranked “full” rankings of analytes that ranked highly in both (1) significance of response to the COCOA intervention (**Table 2**), and (2) significance of association with MPI slope (**Table 3**). A focus on the “full” rankings can identify analytes that have an importance driven primarily by a single or a few individuals. The intersection between these lists is shown in **Table 4** (together with the intersection of the “robust” lists).

Propionylcarnitine is a top personal mediators derived from the “full” analysis but is not found among the top “robust” mediators (identified in lemma 4). This may be type I error. However, if it represents a true mediator, even though significance in our analysis is largely driven by only one or a few participants, propionylcarnitine could represent a mechanistic pathway available to a broader range of individuals. For example, there may be aspects of the intervention that were difficult to comply with, and only benefitted a few individuals able to comply. If we can identify such intervention aspects through further analysis (perhaps on data from future more fully enrolled trials), we may be able to drop compliance barriers or use alternative interventions targeting propionylcarnitine or another yet-to-be-identified personal mediator to broaden the individuals that could leverage one or more of these personal mediators to drive cognitive benefit.

### 3.F. Lemma 6: Potential mediators and mechanisms are not random

Random distribution of mechanistic attributes of potential mediators would be predicted if they all arose due to type I error. Non-random distribution argues against our list of potential mediators all being false positives. However, there could be systematic biases that cluster false positives, so this proof of this lemma alone would not constitute overwhelming proof of our overarching hypotheses. We list all decent analyte mediator candidates from any of the above analyses (**Table 5**). Of the 1461 analytes, 50 had p-values < 0.05 in the “full” analysis. Of the 1461 analytes, 59 had p-values < 0.05 in the “robust” analysis; two of these had p-values < 0.000034 (0.05/1461). We would have expected about 73 analytes to have p < 0.05 if all were due to random type I error, so other than the top two most significant analytes, it is difficult to make a univariate case that any single one of these analytes has a true association with cognitive trajectory. If the co-occurrence of significant intervention and cognition p-values was random, one would expect the directionality of the effect to also be random. However, all eight of the top robust candidate mediators have the same direction of effect, which supports the case that many of these may be true mediators. There seem to be more metabolites in **Table 5** than proteins. But we measured more metabolites, so this may not be significant. Although both types of biomolecules seem to convey some mediation, it is possible that metabolites may play a more important role. Or — they may be easier to detect, as they may be more likely to conduct information via their concentrations, whereas proteins may more likely use other modalities (e.g., covalent modification) for conducting information.

Other than “tryptophan metabolism”, we do not see highly significant overlap between this list of “best” potential mediators and previously curated lists. We surmise that this is because (1) we have identified relatively few mediators, (2) these are distributed across multiple biological entity sets, (3) previously curated biological entity sets typically include only a single class of bioentity (gene or metabolite), and (4) there are no well-curated lists of “biomolecules that mediate cognitive improvement in AD”.

Therefore, to futher our claim that COCOA mediators are not random draws from the list of all possible 1461 studied analytes, we also rely on clustering derived solely from COCOA analyses, and not on previously curated lists. We can illustrate this clustering with a force-directed layout of the correlation network. We graph correlations in **Figure 5**. We consider the 17 top values from lemma 4, the candidates for best robust mediators. If we consider for each of these 17 analytes all 1460 correlations with each of the other analytes, and among those 1460 correlations, identify the rank of the best correlation to another one of the group of 17, the mean rank is 23.6 and the median is 5.5. The expected value under random assortment would be 85.9. Thus, these 17 analytes are surprisingly clustered.

**Figure 5.**
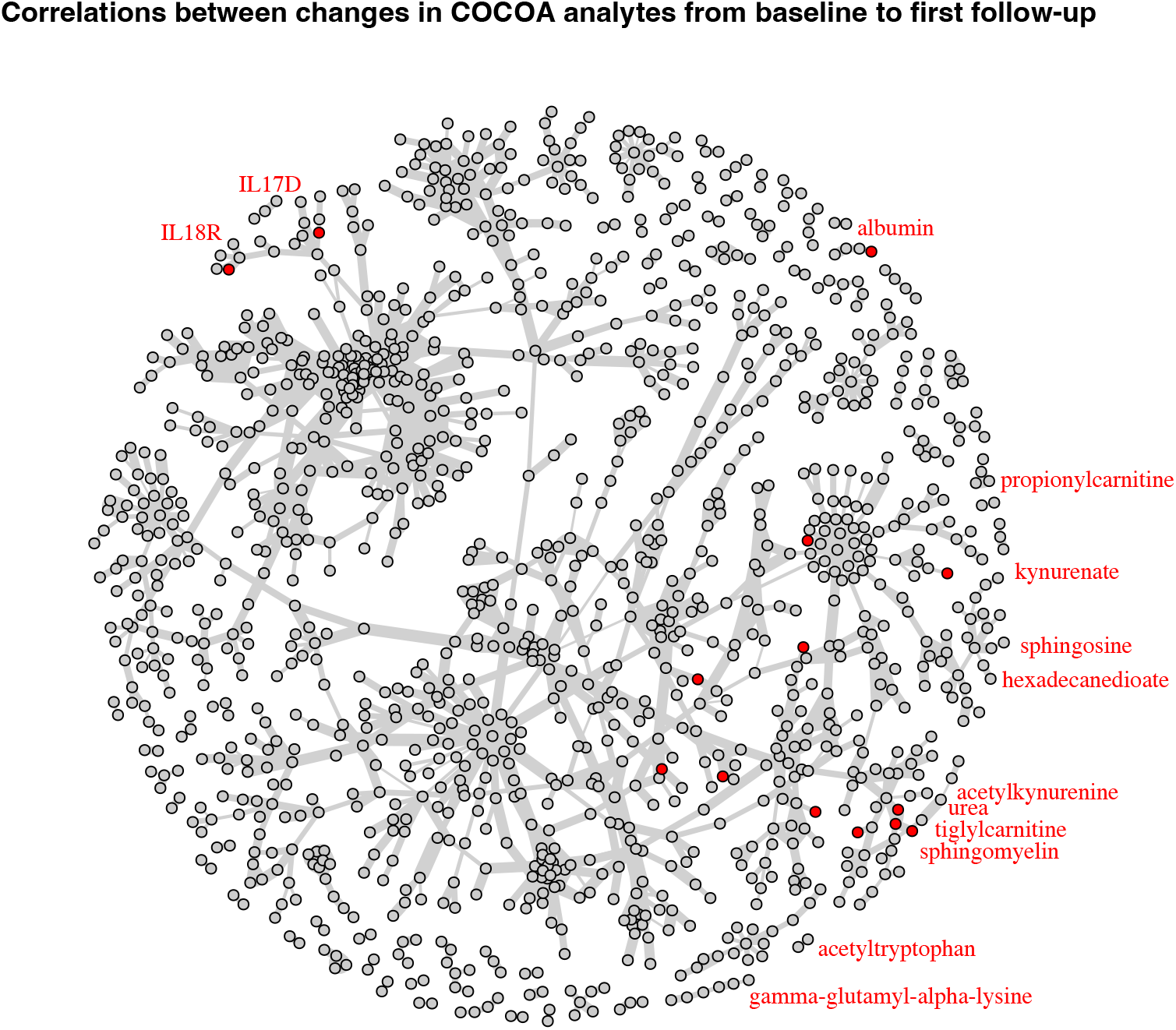
Candidate mediators of the COCOA intervention loosely cluster into two groups. Absolute correlation distances (edges) are shown between highly correlated COCOA analytes (nodes), based on changes between the first and second COCOA timepoints. One cluster including IL17D and IL18R1 may implicate an immune or inflammatory pathway(s) in mediation; another cluster may indicate metabolic pathway(s) in mediation. The non-random distribution of candidate mediators would be unlikely if they were all false positives (noise). This high-correlation network of all analytes is visualized via the Fruchterman-Reingold algorithm (Fruchterman & Reingold, 1991). This network includes the best edge for each node if that edge has a correlation of at least 0.55, and all edges with correlations of at least 0.85. This graph emphasizes the relationships and resulting correlation paths determined by the strongest most significant correlations between analytes, whereas the visualization in **Figure 3** is influenced by subtle effects from hundreds of weak correlations in the all-by-all analysis. Edge thickness ∼ correlation; red indicates candidate mediator (**Table 5**). Labels are shifted radially out from the node they label to avoid obscuring network connectivity. The placement of albumin is arbitrary as it is not highly correlated to any other analyte.

### 3.G. Lemma 7: Several identified mechanisms have considerable prior evidence for mediating AD interventions

There are two lines of prior arguments that suggest that there are likely several — if not many — molecular mediators in the blood that might be identified by the COCOA study. First, many mediators have been suggested in the literature, with varying degrees of evidence (e.g., Niranjan, 2013; Verdile et al., 2015). Although most of these devolve from studies of *causes* of AD, many are also likely to mediate ameliorative treatments, presumably with an opposite direction of effect (although homeostatic/allostatic systems may seek balance and therefore direction of effect may be contextual). Second, since we observe an effect of the COCOA intervention on cognition (lemma #1), there must be *some* mediator(s). Since the main COCOA modalities are diet (including xenobiotics) and exercise, and the vast majority of biological information conveyed by these modalities is conducted through the blood (after transduction through gut, muscle, heart, and other organs), there are likely several if not many mediators of their effects to be found in the blood. These arguments establish a high prior probability that a number of molecular mediators should be identifiable in the blood. These a priori arguments fell short of certainty that COCOA would identify mediators: (1) there could be neurocognitive signals conducted exclusively through the nervous system and brain, (2) there could be signals exclusively conducted by molecular meditators or modalities not assayed by COCOA (e.g., nitric oxide, extracellular vesicles).

One might expect candidate mediators for ameliorating AD to be on a list of “well known suspects”, or closely linked to such suspects in knowledge graphs. For example, one might expect a gene implicated by AD GWAS to have its gene product identified, or an analyte functionally associated with that gene. None of the candidate COCOA mediators intersect with a list of GWAS-implicated genes (Seto et al., 2021). This could be construed as evidence that the list of candidate COCOA mediators is not enriched for true positives and could be entirely composed of false positives. It might also be the case that most mediators of multimodal interventions identified by COCOA target pathways are distinct from the pathways conveying genetic causes of AD. COCOA interventions might target (1) environmental causes of AD, and (2) pathways that do not cause AD, but rather ameliorate AD even when that AD is caused by other pathways. To this end, we examined published evidence that COCOA candidate mediators are in pathways implicated in treatment of ADRD.

#### Kynurenate, N-acetylkynurenine, N-acetyltryptophan

All of these compounds are present in metabolic pathways revolving on tryptophan (KEGG “tryptophan metabolism” module #38; www.genome.jp/module/M00038). Kynurenate is more influenced by KMO (OMIM 603538) than any other gene (**Supplemental Table 2**), and KMO influences N-acetylkynurenine to nearly the same extent. KMO encodes kynurenine 3-monooxygenase (EC 1.14.13.9), which catalyzes the hydroxylation of kynurenine to form 3-hydroxykynurenine. KMO functions at a branching point for tryptophan degradation. Metabolites in the kynurenine pathway are thought to play an important role in neurodegenerative disorders, including AD (OMIM:104300) and Huntington’s disease (OMIM:143100). In these disorders, glutamate receptor-mediated excitotoxicity and free radical formation have been correlated with decreased levels of kynurenic acid. Inhibition of KMO shunts the metabolic pathway towards enhanced production of kynurenic acid, which had been shown to reduce neuronal vulnerability in animal models by inhibiting ionotropic excitatory amino acid receptors and is neuroprotective in animal models of brain ischemia (Zwilling et al., 2011). According to OMIM, these and other findings support a link between tryptophan metabolism in the blood and neurodegeneration. Dietary tryptophan has been suggested as a target for fighting age-related neurodegeneration by precision nutrition (Milošević et al., 2021). Favorable changes in tryptophan metabolites (including kynurenine) together with hexadecanedioate are associated with exercise. These associations differ between men and women. They could at least partially account for sex-dependencies in responses to the COCOA intervention. A caveat to these conclusions is the substantial mean baseline difference between arms of kynurenine (p = 0.0002) and to some extent (p = 0.02) kynurenate. Although not itself a candidate, kynurenine is the most significantly different baseline metabolite, and may serve as a biomarker for other tryptophan pathway bioentitities. Thus, changes in the kynurenine and related pathways could be attributable to regression towards the mean. A further caveat is that tryptophan metabolites may vary as a function of dietary composition, particularly of dairy products, as well as the length of time since the last meal. These factors, which might be highly variable and non-linear over the many months of COCOA, could increase the likelihood of a false mediator attribution to analytes in tryptophan pathways (Solianik et al., 2022).

Most of the identified tryptophan-pathway candidates are identified by the “robust” COCOA analysis. However, kynurenate is only highlighted by the “full” analysis. Kynurenate is a case of an idiosyncratic and nonlinear relationship between analyte change and cognitive change. In these cases, one or a few individuals show a large change in both values, in the context of a smaller aggregate trend across the remaining individuals (**Supplemental Figure 10**).

#### Tiglylcarnitine & propionylcarnitine

Two other analytes that show idiosyncratic relationships identified by the “full” analysis are substrates of HSD17B10 (OMIM 300256). HSD17B10 is also known by other names, including ‘amyloid beta-binding polypeptide’ (ERAB), and ‘amyloid beta-binding alcohol dehydrogenase’ (ABAD). HSD17B10 encodes 17β-hydroxysteroid dehydrogenase X, a mitochondrial enzyme that acts on a wide spectrum of substrates, including neuroactive steroids, isoleucine, and fatty acids, with a preference for short-chain methyl-branched acyl-CoAs (Yin et al., 2022). One of its actions is to inactivate sex steroid hormones (He et al., 1999; Yan et al. 1997). HSD17B10 is present in the postsynaptic density complex and thought to be involved in maintenance of synaptic function (Yang et al., 2007).

HSD17B10 is overexpressed in brains of individuals with AD and MCI; in these brains, substrates including tiglylcarnitine are reduced (He et al., 2018; Costa et al., 2020). He et al. (1999) postulated that neuronal overexpression of HSD17B10 would deplete and thus weaken the protective effects of estrogen, possibly increasing the risk for AD. Supporting this hypothesis, Lustbader et al. (2004) demonstrated that beta-amyloid interacted with HSD17B10 in brain tissue from patients with AD and transgenic mice with mutations in the APP gene. An HSD17B10 peptide inhibited HSD17B10 and β-amyloid interaction and suppressed β-amyloid-induced apoptosis and free radical generation in neurons. Transgenic mice overexpressing HSD17B10 in an amyloid beta-rich environment manifested exaggerated neuronal oxidative stress and impaired memory.

Tiglylcarnitine is more influenced by HSD17B10 than any other gene (**Supplemental Table 2**). Propionylcarnitine and tiglylcarnitine are both acylcarnitines, and substrates for HSD17B10. In COCOA, both of these substrates decrease in response to the intervention, and both are negatively correlated with cognitive amelioration. Our observations could call into question the causality postulated by He at al. For example, instead of causing MCI and AD, overexpression of HSD17B10 might be an allostatic protective response. However, it is more likely that the mode of causality of therapeutic benefit for these two analytes in COCOA does not go through the mutual upstream enzyme HSD17B10, but rather is a direct effect of reducing these metabolites (and their precursors) via dietary modification.

A precursor to propionylcarnitine, propionate, is thought to mediate neurotoxicity via impairment of the urea cycle (also implicated by our COCOA data), citric acid cycle, respiratory chain complex, and lycine cleavage subsystems (Killingsworth et al., 2021). L-carnitine deficiency may potentiate propionate-mediated neurotoxicity by and preventing the conversion of propionyl-CoA into propionylcarnitine (Roe et al., 1984; Maldonado et al., 2016). Varma et al. (2018) associate lower blood concentrations of propionylcarnitine with lower CSF levels of Aβ1–42 in a preclinical AD cohort, suggesting a connection between propionylcarnitine and amyloid metabolism in individuals along the AD spectrum.

Acylcarnitine concentrations change in blood and urine in response to changes in dietary fat and protein (Khodorova et al., 2021). In particular, the short-chained acylcarnitines including propionylcarnitine and tiglylcarnitine are associated with changes in high-fat diets. Most likely, COCOA participants decreased their consumption of high-fat diets in response to coaching while controls did not. Tiglylcarnitine and propionylcarnitine are reduced following ketogenic diet (Cappuccio et al., 2017). COCOA coaching aims to personalize diets primarily along nutrition recommendations such as those based on the DASH dietary pattern or MIND Diet (Roach, Hara, Fridman, et al., 2022), which both promote low fat and low calories, so would be expected to reduce short-chain acylcarnitines in COCOA coached participants compared to COCOA control participants. Such reductions were indeed seen in COCOA.

#### Albumin and urea

Albumin and urea are particularly interesting because: (1) they very commonly measured as part of routine clinical labs — urea is one of eight assays in a typical basic metabolic panel, (2) they are very inexpensive to measure, (3) they are part of nitrogen metabolism pathways implicated by other significant mediator candidates, and (4) urea has been previously associated with AD (Ramdane, 2017; Chen et al., 2021). Albumin changes — but possibly in the opposite effect direction — have been reported to impact aging and neurodegeneration. Low serum albumin is associated with increased odds of cognitive impairment (Llewellyn et al., 2010). Albumin oxidation is higher in AD than healthy controls (Costa et al., 2018). Albumin exchange has been proposed as a treatment for AD (Menendez-Gonzalez & Gasparovic, 2019). Working out the directionality of causal influences could help explain the opposite effect directions of these studies and COCOA. One possibility is that both very high and very low albumin are detrimental; most Americans — probably including the baseline COCOA population — consume too much, so the aggregate effect in COCOA is beneficial for albumin reduction.

If many other identified biomarkers from our effort at least currently require somewhat expensive assays, urea trajectory alone might be immediately employable inexpensively as a reference for personalization of therapy in nearly all settings. Albumin, with higher significance than urea, may be a similarly accessible metabolite that also reflects nitrogen and overall protein flux. Babygirija et al. (2022) report beneficial effects of protein restriction on metabolic health, cognitive function, and neuropathology in a mouse model of Alzheimer’s disease (see also Babygirija & Lamming, 2021). Tynkkynen et al. (2018) note an inverse correlation between creatinine and dementia risk, which they hypothesize could be related to muscle mass, and could also be linked to diet. Not all studies have replicated this particular relationship, perhaps because of differences in populations and environments, but Tynkkynen et al.’s findings support a potentially complex interplay between nitrogen metabolism and ADRD risk. In COCOA: albumin, urea, and other components of nitrogen metabolism may be reflecting the importance of the nitrogen metabolic subsystem. A caveat to these conclusions is the mean baseline difference between COCOA arms of urea (p = 0.006); changes in urea could be attributable to regression towards the mean.

#### Hexadecanedioate

Hexadecanedioate is influenced by exercise. The reportedly beneficial direction of effect of hexadecanedioate is mixed. Nayor et al. (2020) report that increases (which is what is seen in COCOA) are beneficial for overall cardiometabolic health, particularly in women. Menni et al. (2015) report increases associated with increases in blood pressure. Sun et al. (2019) report increases associated with increased stroke risk, possibly due to increased blood pressure. Chen et al. (2021) see a negative correlation with AD but a positive correlation with frontotemporal dementia (FTD). The relationship of hexadecanedioate with AD amelioration may be important, complex, and non-linear. In combination with other biomarkers and systems modeling, hexadecanedioate may become important in guiding personalizing interventions, as an intervention that alters hexadecanedioate beneficially in one individual may be detrimental in another.

Hexadecanedioate is related to biosynthesis of unsaturated fatty acids (KEGG pathway #73; www.genome.jp/pathway/map00073). As mentioned above in conjunction with tryptophan metabolites, hexadecanedioate is also increased with exercise (possibly differently between sexes) and promotes cardiovascular health and lower blood pressure (Menni et al., 2015). Exercise recommendations, perhaps coupled with diet recommendations in COCOA may promote changes in these mediators.

#### IL18R1 and IL17D

Inflammation may play a role in causing or potentiating ADRD (Leng & Edison, 2021). Two of the candidate mediators for the effect of the COCOA intervention are proteins that mediate inflammation in other contexts, so may also mediate a protective effect due to the COCOA intervention. The IL17 cytokine family functions in inflammation and autoimmunity, with most of the family members expressed in Th17 cells (Li et al., 2019). IL17D is expressed broadly in nonimmune cells (Liu et al., 2020). IL17D is regulated by nuclear factor erythroid-derived 2–like 2 (NFE2L2, aka Nrf2), an oxidative stress sensing transcription factor that binds to antioxidant response elements (AREs) in promoters of genes (Saddawi-Konefka et al., 2016). Activation of NFE2L2 target genes in astrocytes and neurons is strongly protective against inflammation, oxidative damage, and cell death (Hannan et al., 2020; Scapagnini et al., 2011).

IL18 plays roles in infectious, metabolic, and inflammatory diseases. IL18 binds to the IL-18 alpha chain (IL18R1, aka IL18Rα), with low affinity, and binds to IL-18 receptor beta chain (IL18RAP, aka IL-18Rβ) with high affinity. Most cells express IL18R1. IL18RAP is primarily expressed on T-cells and dendritic cells. IL18, through its receptors, activates inflammation pathways, including those mediated by NFκB (Kaplanski, 2018).

#### Palmitoyl-sphingosine-phosphoethanolamine & sphingomyelin

These metabolites are related to sphingolipid metabolism (KEGG pathway #600; www.genome.jp/pathway/map00600). Sphingolipids are implicated in neurodegeneration (e.g., Alaamery et al., 2021; Varma et al., 2018). Palmitoyl-sphingosine-phosphoethanolamine is more influenced by PCSK9 than any other gene (**Supplemental Table 2**). PCSK9 may influence AD (Mazura et al., 2022; Picard et al., 2019). Although not all lines of evidence point to PCSK9 as being a cause of AD (Paquette et al., 2018; Courtemanche et al., 2018), it may still be part of a targetable subsystem to treat AD, as suggested by Abuelezz et al. (2021). Shippy et al. (2020) also observe correlations between dietary interventions, changes in sphingomyelin analytes, and AD.

#### Aspects of the COCOA intervention may harm

Given the complexity of the COCOA intervention, it is reasonable to hypothesize that some aspects of it may harm. Given the overall beneficial outcome, if these harmful mediations are occurring, then they are outweighed by mediators that ameliorate. At least one metabolite (gamma-glutamyl-alpha-lysine) meets our criteria for mediating an effect of the COCOA intervention on cognition, but with the direction of the effect harm rather than amelioration. This could be a false positive; the probability of a false positive is enhanced by a slight difference between arms of this analyte at baseline (p = 0.013). There is little literature specifically on gamma-glutamyl-alpha-lysine. However, there is considerable literature on the class of compounds encompassing gamma-glutamyl-alpha-lysine: the gamma-glutamyl amino acids. For example, Wang et al. (2020) report that gamma-glutamyl amino acids including gamma-glutamyl-alpha-lysine play a role in AD risk, particularly in the female brain. It might be possible to revise an intervention to remove the aspects that harm (e.g., reduce gamma-glutamyl-alpha-lysine) or, if not causal, the true causal harmful correlates. On the other hand, a reduction in a particular harmful mediator may be inextricably linked to beneficial mediators.

#### Some aspects of the COCOA intervention may cause substantial systemic effects, but not be related to cognitive amelioration

We expect most changes in response to diet, exercise, and the other COCOA interventions to <u>not</u> have cognitive effects. For example, 3-formylindole is significantly decreased in the intervention arm but has no significant effect on MPI slope. This metabolite is derived from dietary tryptophan and the gut microbiome, so is expected to change following dietary interventions. Other highly significant analytes, such as 1-palmitoyl-2-oleoyl-GPI, are also known to change in response to diet (e.g., Jewell et al., 2020). The COCOA intervention may have benefits to health unrelated to cognitive amelioration, mediated by such analytes. Some of these benefits (e.g., stroke prevention), if they exist, may affect cognition indirectly and might be detectable in a longer or larger study. Given the known response of 3-formylindole to dietary interventions, and our result of 3-formylindole as the most significantly changing analyte (“robust” analysis) in response to the COCOA intervention, we gain further confidence that our list of most significant analytes contains at least some true positives.

#### Some analytes associated with cognitive amelioration may not be related to the COCOA intervention

A wide variety of individuals participated in COCOA and pursued diverse lives independently of each other. Differences in genetics and environment could lead subgroups of participants independent of arm to share a mediator of cognitive effect. For example, CD163 is a microglial marker associated with AD (Roberts et al., 2004; Pey et al., 2014). It is significantly associated with MPI slope in COCOA, but not with the COCOA intervention. Other analytes with similar significances of association are gamma-glutamylglycine and gentisate. Such analytes might mediate (or reflect) the effects of environmental insults such as episodes of infectious disease or stress uncorrelated with the intervention. They might also mediate the effects of beneficial non-coached environmental factors, or genetic factors.

#### Subsystems

Chen et al. leveraged two-sample Mendelian randomization and GWAS summary statistics to explore the causal association between 486 metabolites and ?ve neurodegenerative diseases (NDDs) including AD (Chen et al., 2021). They suggestively associated 164 metabolites with the risk of at least one neurodegenerative disease. They found metabolic pathways involved in NDDs, including “urea cycle”, “arginine biosynthesis”, “purine metabolism”, and “D-glutamine and D-glutamate metabolism” for AD. They also found “phenylalanine, tyrosine and tryptophan biosynthesis” for ALS and MS. “Carnitine synthesis” was associated with FTD. Leao et al. (2021) show that the acylcarnitine subsystem differs between controls and AD patients. These pathways markedly overlap the pathways implicated by our current analysis. Particular metabolites implicated in AD included urea and hexadecanedioate. Hexadecanedioate is also implicated in ischemic stroke (Sun et al., 2019), possibly mediated by its effects on blood pressure and/or its hypolipidemic, anti-obesity, or anti-diabetogenic properties. These pathways overlap pathways implicated in ADRD, including pathways affected by APOE, as well as potentiation of AD by a vascular dementia diasthesis.

#### Connectivity of AD

Essentially all of the top candidates for mediators are plausibly connected to AD. This could be considered evidence that these candidates are true positives and represent excellent biomarkers and clinical targets. On the other hand, it may be the case that AD is so well studied that the literature is so replete with false or misleading positives that almost any analyte would be found to be related to AD in manners similar to those described for the candidates above. For example, hypotaurine is a metabolite near the bottom of our candidate list; at least two articles in PubMed include both ‘hypotaurine’ and ‘Alzheimer’ in their Abstracts and could be plausibly linked to AD using arguments similar to the arguments for consistency that we present above, although perhaps not as strongly. There are methods of quantifying the surprise, or significance of connectivity of set of bioentities to AD, typically operating on knowledge graphs such as underpin Translator (Wood et al., 2022). These methods require some simplifications, as they are currently not able to capture all nuances of relationship; we defer such objective analysis to a subsequent manuscript. In the meantime, we subjectively judge our level of consistency surprise to be moderately but not absolutely convincing.

## 4. Discussion

We have shown that personalized multimodal lifestyle interventions can ameliorate cognitive trajectories in individuals on the AD spectrum. These interventions cause many changes in blood analytes. Several of these analytes are also correlated with cognitive amelioration. Analytes that are altered following the intervention ***and*** that are correlated with cognitive amelioration are candidate mediators. A mediator mechanistically (causally) conveys the influence of the intervention, propagating its information through biological pathways, to the brain, where it influences the final common pathways of ADRD pathophysiology, presumably on neurons and their synapses. COCOA was not designed to detect *all* possible mediators. The mediators identified by COCOA implicate inflammatory and metabolic pathways that respond to changes in diet and possibly exercise.

Our results have a number of broad implications. First, COCOA served well as a pilot trial. More trials should be modeled after COCOA’s trial design. Such designs can generate new information, both testing well-defined hypotheses and expanding the scope of inquiry into areas of pathophysiology that are less explored. Second, ADRD can be treated using mediators that are not directly related to the most prominent pathways (e.g., amyloid hypothesis) targeted by many pharmaceutics. Pathways to *treat* ADRD may be different than pathways that *cause* ADRD. Lifestyle treatments are likely to complement, not replace, pharmaceutical treatments. Both are likely to be part of many optimal personalized multimodal therapies. Third, lifestyle interventions should be emphasized in current treatment for all individuals on the ADRD spectrum. Even without the additional evidence from the COCOA trial, there is strong evidence to elevate the prominence of these recommendations in guidelines and provider training. Lifestyle recommendations are often buried as afterthoughts at the end of multipage clinical recommendations that otherwise focus on pharmaceuticals (e.g., Alzheimer’s Association, 2019). Lifestyle recommendations often lack specifics, making them hard to implement, particularly for practitioners who are not experts in neurodegeneration. Such recommendations should be placed before pharmaceutical recommendations in professional clinical guidelines and given equal amounts of text / importance. More research — some of which is ongoing, such as POINTER — is necessary to make this case even more convincing (Coon & Gómez-Morales, 2022). More funding for lifestyle interventions should be applied both to research and via payers for medical care.

### Nonlinear responses

Many — perhaps most — published AD interventions have nonlinear responses (e.g., Burns et al., 2007; Raskind et al., 2000) and most occur in the first six months to a year following the start of the intervention. Therefore, we may have been overly simplistic in proposing a linear hypothesis as foundation for our primary outcome test. However, it would have been difficult without much more preliminary data to choose an appropriately powered nonlinear model a priori. More pilot studies and trials like COCOA may enable improved trial design, including picking better, nonlinear models for assessing primary trial endpoints.

### Two-year study

COCOA maintains its effect for at least a year. This is not always true with pharmaceutical interventions, but similar plateaus have been seen with multimodal interventions (e.g., FINGER). Many published pharmaceutical trials end after six months to a year; stakeholders have a disincentive to test a drug longer than 6 months. Donepezil was approved by the FDA based largely on 6-month data; any longer trial of a new drug might risk showing inferiority. Despite this, a few trials of small-molecule pharmaceuticals have extended as long as 2 years (e.g., Bullock et al., 2005). These have established limited benefits for some small molecules (Herrmann et al., 2013). Trials of monoclonal antibodies have typically lasted longer than small-molecule trials. Notably, the DIAN trial for dominantly inherited AD lasted up to seven years (but the intervention did not slow cognitive decline) (Salloway et al., 2021). These trials have recognized the benefit of relatively early intervention (Aisen et al., 2022) and the possibility that transient non-linear effects might fade after a few months. To avoid distraction by transient effects, comparative reviews and meta-analyses of intervention trials for AD should focus on trials lasting at least 12 months; COCOA was designed to enable inclusion in such comparisons.

### Individual/idiosyncratic responses

We observed a number of candidate molecular mediators (e.g., propionylcarnitine) with their best evidence stemming from only one or a couple of participants. These would need to be seen again in other studies to validate any of their utility. Even though rare in COCOA, they may ultimately be more broadly applicable. Even though they did not appreciably leverage these rare mediators in COCOA, other COCOA participants might under some circumstances benefit from altering that mediator. Some of these “rare” mediators may actually bridge the spectrum of effectiveness. Some analytes seem to convey a big effect in a few individuals but also convey slight effects in many individuals (e.g., **Supplemental Figure 10**). There may be an opportunity to discover better ways to target coaching and specific interventions to specific people to enable them to change that mediator and improve their AD trajectory. Ideally, to fully translate such an insight, the causative factor and underlying pathology addressed by that mediator could be identified, so that interventions targeting that mediator are targeted to the optimal subset of the population. Some mediators might be so rarely used that almost all knowledge of that mediator would have to come from the same person who would benefit from that knowledge. This would require frequent assays to establish the trajectory of that mediator and very sophisticated mechanistic models to judge the appropriateness of that mediator and to guide therapy. In clinical practice, even if this extreme is never reached, it is likely that the best molecular blood biomarkers for monitoring response to AD therapy will be different for different patients.

Many of our identified mediators have stronger and/or more significant effects from the intervention than they may have for their correlation with cognitive outcome. This is expected: we expect to see big effects of diet and exercise on blood analytes. Although there are some analytes that could – in theory – cause big changes in cognition, such as the uremic toxins, we don’t expect these particular analytes to be in play in any other than a minor way due to the COCOA intervention. Rather, we expect a number of small effects of analyte changes correlated with cognitive changes. The example in **Supplemental Figure 10** may be typical of the expected strengths of effect sizes likely to emerge from the workflow presented here in the context of COCOA data. Some or even all of these may not be directly casual, and merely reflect causality of correlated bioentities, and so may have muted effect sizes. Conceivably these analytes could have substantial effects if they were better or more aggressively targeted.

### There are numerous limitations to the COCOA trial

Our results may not be generalizable to other populations. COCOA participants have mostly self-described as White. However, it is increasingly apparent, not only in the context of AD and dementia, but also throughout all of human biology, that the amount of intra-individual variation in humans tends to be greater than the amount of intra-group variation, with the exception of a few very carefully chosen definitions of ‘group’ (e.g., Rosenberg et al., 2002; Yatsunenko et al., 2012). It is therefore important to design trials that measure as many variables as possible to best understand the diversity represented in the studied population. As a pilot study, COCOA has demonstrated the feasibility of including many diverse individuals in trials designed with dense data collection and analyzed with systems epistemology. We conclude that more resources should be devoted to fund larger multi-center dense-data trials — not only of AD but also other diseases — that can be inclusive of the full diversity of the global population. To this end, inclusion and exclusion criteria should be made as loose as other trial-design constraints allow.

In many instances we use conservative statistical approaches and parameterizations to facilitate high-throughput analyses uniformly across all analytes. For example, we apply sex adjustment to all regressions. Such adjustment is unnecessary in many cases and reduces power by unnecessarily adding one parameter estimation. Likewise, unless otherwise mentioned, all p-values are two-sided. Many analytes have substantial prior knowledge that would argue for a one-sided test for that analyte. Thus, for many analytes, we are underestimating significance.

Differences between participants at baseline are a limitation of COCOA and all other clinical trials. Because these differences are also a strength as well as a limitation, it may be unreasonable to attempt to eliminate them completely in trial designs, even if it were possible. These differences will result in differences between baseline distributions of variables. These differences could have two consequences for interpretation of trial results. First, such differences — and not the intervention — could be responsible for the difference in primary outcome between arms. Second, such differences may result in regression toward the mean of particular variables, and lead to false inferences of correlation and causality. In future analyses of COCOA data, we plan to include more timepoints and/or consider analytes in the context of external reference distributions to better identify and quantify effects of regression toward the mean. One particular concern would be a difference in mean baseline MPI score (**Table 1**). Although not significant, there is a subtle difference. However, a higher baseline MPI score in the control compared to intervention arm would be predicted to result in less MPI loss, since the rate of cognitive decline tends to accelerate as a function of itself. Since we observe the opposite, we doubt this baseline MPI difference had any impact on our overall conclusions. If anything, it might suggest the effect of the intervention is slightly stronger than we report. Attrition from a trial, whether biased by the intervention or not, can also result in differences between outcome and other variables. We did not detect any significant differences in attributes of individuals withdrawing from COCOA (Roach, Hara, Edens, et al., 2022). In particular, neither (1) arm, nor (2) MPI score at the time of withdrawal had a significant impact on retention.

It is possible but unlikely that poor randomization could have affected COCOA. For example, if individuals with a propensity to self-motivate and seek out life-changing behaviors such as dieting were overrepresented in one arm, we could falsely attribute the original source of the effective intervention to coaching rather than to individual inspiration. A poor randomization is unlikely for three reasons: (1) we cannot identify any aspect of our methodology that would have biased randomization, (2) baseline demographic variables are evenly distributed (**Table 1**) and (3) we clearly see systemic changes due to the COCOA intervention (**Figure 3 & 4**). These changes broadly impact multiple molecular subsystems. So any false rejection of a null hypothesis due to unfortunate assortment at the randomization stage would have had to create a biased set of people not only instantaneously different at baseline, but also who just happened to be primed to differently undergo change across a set of analytes consistent with expectations from prior research/knowledge. However, even if poor randomization defeated the epistemology of arm assignment, the remaining conclusions drawn from the cohort-oriented and individual level analyses would hold.

Another limitation of COCOA is that — despite our efforts to assay “everything” — not all analytes are assayed. At the time of trial design, cost constraints limited the trial to assaying only 443 proteins. Today, it would be more feasible and less expensive to assay thousands of proteins encompassing more systems (e.g., endocrine) and provide more thorough exploration of others (e.g., immune). We measure only metabolites that are measured by Metabolon’s global metabolite analysis service that employs mass spectrometry. Very small or volatile metabolites such as nitric oxide are missed, for example. Any metabolite present at trace amounts, below threshold of detection for our assays would be missed. Also, some assayed analytes may be meaningfully altered, but these alterations may not be appreciated. Variability due to measurement error could reduce our power to detect a change. Inherent biological variability such as periodicity may not be appreciated by our analyses that focus on long-term baseline changes. These periodicities could include a short-term or one-time pulsatile effect with profound system implications, followed by a return-to-baseline of that analyte. Or they could be reflected by a change in period of a cycling analyte. Unappreciated and uncompensated external confounders could obscure the impact of some analytes. For example, phlebotomy conditions can influence blood analyte measurements (Devi et al., 2021); this particular confounder is unlikely due to standardized COCOA protocols but exemplifies one of many subtle effects that might not be appreciated. Lastly, some analytes may convey causal information without being altered in amount or concentration. The design of COCOA and other dense data trials may allow for lessened impact of such confounders, through increased ability to detect and appreciate their influence. For example, we can measure caffeine metabolites, so if caffeine were a suspected confounder, its effects could be modeled. Another class of unappreciated analytes are those that change their concentrations in locales other than the blood (or in unresolved subfractions of the blood). Some proteins may be modified in ways that are transparent to the Olink assay technology. Isomers may not be resolved by mass spec.

Yet another limitation of COCOA is outcome measurement variability. In the context of high variability, aggregate results are not necessarily applicable to any given individual (**Supplemental Figure 5**). Such variability underscores the importance of personalized analyses to understand these very different individual trajectories. Cognitive outcome measures with less error are a general need for studies of neurodegeneration and further development of such measures should be a research funding priority. Also, since cognitive performance can fluctuate over the course of a day or any short time period, studies will also benefit from frequent if not nearly continuous assessment to better power tests of long-term change in cognitive baseline (see also Roach, Hara, Edens, et al., 2022).

At the outset of COCOA, we were skeptical that we would see any effect of the intervention or identify any mediators. This skepticism increased as event-driven modifications materialized. However, as a result of the data analyses presented here, we are at least moderately convinced that there is an effect and that we have identified some mediators. The biological plausibility of the emergent mechanisms carries considerable probative value. For example, we are likely to change our own personal lifestyles in light of these conclusions. Participants and clinicians participating in the trial — by the time the trial was over — were increasingly convinced based on their subjective and objective experiences with COCOA that the intervention was beneficial and sought for ways to continue with the COCOA intervention after their formal participation ended. The fact that a strong case for lifestyle interventions can be made on a trial that was at the outset intuitively underpowered (at least to some) suggests three somewhat unrelated conclusions: (1) there is a much bigger effect of lifestyle interventions on AD trajectory than we believed, (2) the mechanisms of that effect are not previously published “above-the-fold headline” mechanisms but rather less-well-known previously published mechanisms, and (3) dense dynamic data trials can extract much more epistemological value from a trial than pauci-data trials.

There are several reasons that COCOA may not have identified all — or perhaps even any — of the mediators, if they exist, of cognitive amelioration following multimodal intervention. COCOA may have been underpowered, at least with respect to some interventions and mediators. COCOA may not have included an effective intervention. Participants may not have been compliant with interventions. COCOA may not have measured a particular mediator, or even any analyte correlated with that mediator. Nevertheless, we have identified some mediators with some confidence. These are not necessarily the most important mediators nor all the mediators. They do serve to bolster — and perhaps even prove — our overarching hypotheses. Multimodal interventions are effective in ameliorating cognitive and functional decline in the early stages of AD. Multimodal interventions work through a variety of mechanisms, suggesting that AD is indeed caused by dysfunction in multiple subsystems and can be ameliorated by improving the function of multiple subsystems. Furthermore, these subsystems differ between individuals and therefore treatment is best personalized. Personalized therapy may best include modalities that are common to many other persons’ best therapies, but also include some therapies guided to very personal circumstances.

Correlation is not always causation. Correlation due to prospective randomization in an RCT is a very strong argument for causation. Thus we can be somewhat confident that the COCOA intervention causes broad molecular changes in the blood, and that the COCOA intervention improves the cognitive trajectory of AD. Although we provide some evidence for specific blood analytes that mediate this causation, there are alternative explanations for mediation. Significance, coherence & consistency (aka, biological plausibility), and effect size alone do necessarily not prove causation. We may be able to conduct further analyses on future COCOA data releases, including time dependency analyses to bolster our case for causation. In the meantime, the possibility remains that participants in COCOA, in addition to altering their blood metabolites, may also be altering other components of their physiology including neurocognitive functions not reflected by blood analytes.

A common upstream cause of both cognitive improvement and of blood metabolic change is possible, without the blood metabolic changes being causative of cognitive improvement. Even if there are parallel causal pathways, we doubt that blood metabolic changes play <u>no</u> causal role whatsoever. COCOA’s evidence as well as accumulating evidence from many other completed and ongoing studies of lifestyle interventions for ADRD are very compelling. Furthermore, even if our proposed blood mediators are not causal, they are likely to reflect useful causal interventions, and therefore can be used (if validated) to guide therapy. They can also be used to further identify and personalize the components of multimodal interventions that are most effective. As more trials like COCOA are conducted, iterations will improve both the utility and the understanding of causality and mechanism for both molecular and non-molecular mediators of lifestyle interventions.

### Comparisons with other case reports, studies and trials

The general utility of multimodal interventions to ADRD is now widely accepted, as opposed to a few years ago — when greater skepticism prevailed (Hendrix et al., 2022). Although the general utility is widely accepted, there are plenty of areas of uncertainty, skepticism, and/or controversy. These areas generally encompass uncertainty about exactly which modes should be included, and the details of the included modes. One area of uncertainty: should cognitive training be included, and if it should, what specific exercises and regimen should be included? Another: although it is generally accepted that diet is important, exactly which diet is best for each individual, and what supplements if any should be included? Given the increasing acceptance of multimodal interventions — to the point where they may now be considered to be the most effective intervention for ADRD — we do not in this manuscript thoroughly review the literature. The FINGER trial is the most recognized trial demonstrating the effectiveness of multimodal interventions (Ngandu et al, 2022). More recent studies, including the Chicago Health and Aging Project (CHAP) and the Rush Memory and Aging Project (MAP), validate these findings (e.g., Dhana et al., 2020). Several other case reports and case series similarly support the importance and effect size of multimodal interventions (e.g., Ross et al., 2021; Toups et al., 2022). Even dietary interventions alone have been reported with similar effect sizes, although not all diets & studies are equal (Kocatürk et al., 2022). Neither the significance or effect size of the COCOA multimodal intervention should be surprising in light of these reports. The COCOA effect size is similar to those observed in other studies and is larger than we anticipated for the purpose of power calculations when we designed COCOA (in 2017). Although the COCOA results presented here do not resolve all the uncertainties of multimodal therapies, they may guide improvements to be used in future iterations of these therapies and demonstrate a research methodology that can be applied iteratively to continue such improvements in knowledge and therapies in the years to come.

### Clinical Insights

A number of candidate mediators (**Tables 4 & 5**) are anticorrelated with cognitive improvement. Reductions in blood levels of these mediators appear beneficial to cognition. Several of these are associated with food consumption, particularly highlighting nitrogen metabolism. Therefore reducing caloric intake, possibly including protein restriction, may be a beneficial component of multimodal interventions in some individuals. Protein or caloric restriction, particularly in the elderly, can be dangerous; interventions must be personalized with an emphasis on harm reduction.

Many if not most dementia patients — including those on trajectories heading towards dementia — in the United States are not receiving the best current standard of care. Standard-of-care includes screening for and treating known causes of dementia, optimizing diet and exercise, and cognitive or social engagement (e.g., Baumgart et al., 2015). ADNI participants probably receive better care than average, as they are recruited to and participate in an academic clinical study. However, they are unlikely to all receive care at specialty memory clinics and may not consistently receive care consistent with modern guidelines, particularly as such care is typically not fully reimbursed by payers. It is possible that even the specialty memory clinics providing standard of care to COCOA participants are not fully following modern recommendations. In that case, one possible interpretation of the beneficial result of the COCOA intervention is that COCOA is “merely” doing a better job of implementing standard of care than is usual. If so, our analysis points urgently to the need to better implement standard of care, and to better emphasize multimodal lifestyle interventions in training and published guidelines.

### Lessons for trial design

COCOA is a pilot study. COCOA explores new approaches to trial design. We have implemented this design for AD. Our experience offers lessons, some of which are generalizable to many situations, and other more narrowly applicable to ADRD. Despite our efforts to generate dense longitudinal data, more and better analyses would be possible if we had more frequent — or continuously measured — outcome variables. Because our outcome was non-linear and much change happened rapidly, we would have liked to have more frequent measurements during the first few months of the trial. As assays become more frequent, they will need to be less burdensome to both staff and participants.

The regulatory scope to implement unusual trial designs is constraining. Most clinical trials are designed to achieve FDA approval for a product. Unusual trial designs invite increased scrutiny and skepticism. Such scrutiny and skepticism is an unreasonable risk to add to other risks undertaken by shareholders and directors of for-profit companies. Since most funding for trials comes from for-profit companies, the vast majority of experts in clinical trial design, funding, and implementation are trained and experienced in conventional trial design and view innovation askance. We do not object in principle to this conservatism. It serves the public well and has created a pipeline of safe and effective pharmaceuticals. However, design of an innovative trial faces many roadblocks at all stages – from inspiration to execution – due to lack of enthusiasm or obstruction from bureaucratic systems. These obstacles range from simple (deposition requirements in clinicaltrials.gov geared towards FDA-style trials), to moderate (requirements by manuscript and grant reviewers to abide by rigid guidelines), to the profound (inability to find funding for innovation). Such obstacles are often justified by momentum: “you cannot do it your way because it is not the way it has always been done”. Aspects of the global environment for clinical trials should change to better accommodate and welcome innovative trials.

Part of the design of the COCOA project was to generate dense molecular longitudinal data on an AD cohort. These data are useful even if the cohort is not participating in a RCT. Much can be revealed about the pathophysiology of a disease from cohort analysis. Correlations between analytes and outcomes are useful building blocks of knowledge even if these correlations are not related to an arm or intervention. All clinical trial designs should elevate the emphasis of cohort data analyses to expand knowledge.

### Future directions

This analysis represents a subset of all possible analyses that could be performed on COCOA data. This subset (data freeze 1) focuses on the first year of blood analyte data of trial participants. Future analyses will focus on subsequent data freezes and include additional data types. Future analyses will also include additional algorithmic approaches and analytical tools. It may also be possible to use the results of COCOA to help modify and refine the best approach for practical clinical recommendations for individuals with or at risk for AD, and to design implementations of recommendations for future clinical trials that will yield the most information about how to further improve clinical recommendations and advance basic biomedical understanding of AD. The PREVENTION trial is in progress and should serve to iterate, validate, refine, and extend conclusions from COCOA (McEwen et al., 2021).

One future direction for analyses of these data is to perform stratified or subgroup analyses. There is a (purely theoretical) philosophical tension between two extreme philosophies, both of which are systems biology strategies. The first is that every individual should be analyzed individually in an “N of 1” analysis. Given sufficient prior and revealed knowledge of the mechanisms of the system, there is no need to perform any aggregate analyses, and all analyzes should be performed on single individuals. This would be the extreme of stratification. The other is to recognize that all individuals, although differing in some respects, share commonalities of systems. Since stratification loses information stemming from these commonalities, any stratification should be avoided. This would be the extreme of aggregation. However, if a subgroup has markedly different system behaviors from other individuals in the population but markedly common behaviors within the subgroup, then a balanced approach between these extreme systems philosophies may be warranted. Stratification is most informative when each subgroup has large enough membership to power classical aggregate statistical approaches. Since the overall number of participants (N_p_) in COCOA ranges from 42 individuals with at least two full omics data sets to 55 randomized individuals, any stratification imperils this aggregate statistical power. Therefore in this manuscript — where we have stratified — we have focused on cases of extreme stratification (N of 1), and leveraged biological plausibility for knowledge extraction. Future trials should consider the desirability of stratified analyses when planning for N_p_ in trial design and implementations. Future COCOA analyses will include stratified analyses, particularly focusing on stratification by sex. Since sex has a significant impact on MPI trajectory, and there is substantial prior knowledge on the systems differences between males and females, such stratification may yield insight.

### Biological plausibility & the rising importance of epistemology for the future of clinical trials

An important component of systems epistemology is *biological plausibility:* placing current research coherently and consistently in the context of previous research. More generally, new observations should be interpreted in the light of previous observations. Much of the argument we present today would not have been as strong a few years ago. In particular, at the time of trial design prior to beginning enrollment of participants, we could not have known the best methods of analysis or computed an exact power for the trial. These uncertainties create practical challenges for launching trials designed to take advantage of future knowledge, because many stakeholders assume that knowledge will be fixed over the course of the trial, including: funding agencies, institutional review boards, trial-design repositories (e.g., clinicaltrials.gov), standardized guidelines (e.g., www.equator-network.org), and journal reviewers and editors. To convince some stakeholders, arguments such as the “Three Billy Goats Gruff” argument (“Trust me, a bigger better analysis will come along soon.”) and the “Stone Soup” argument (“This analysis will be much better once I add in other people’s knowledge.”) must be made convincing. One way to do this is empirically, by demonstrating successful execution and knowledge generation from trials such as COCOA. Additionally, recognition of trends in epistemology such as the exponential increase in knowledge of blood metabolites can aid proselytization. Although not used extensively in our COCOA analyses to date, artificial intelligence (AI), particularly hyperscale AI, is likely to potentiate the value of dense-data studies (Bughin et al., 2017; Naylor, 2017).

Our current analyses to support our overall claim focus on strength, correlation, and biological plausibility. In this case, biological plausibility includes systems analyses of newly generated data coupled with prior (published) knowledge (Roach, Hara, Fridman, et al., 2022). It is not necessarily for our syllogism that we explicitly identify any or all components of the causality pathway linking interventions to outcomes. However, such identifications advance the coherence and plausibility of our argument. They also advance basic biological understanding of AD, one of the goals of the COCOA project.

Consistency and coherence increase in value the more prior knowledge is available and the more dense data is available for a trial. These factors of epistemological evidence are already in common use: reviewers of clinical science often reject claims based on strong significance and effect size unless such claims are also biologically plausible — that have a mechanism. The existence of such a mechanism adds probative value. The overall cohesiveness of our data increases our confidence (beyond mere p-value) that the COCOA multimodal intervention, in aggregate and on average, ameliorates cognitive decline in early-stage AD. However, it is currently hard to quantify this increase in confidence. It is a moderate but not overwhelming increase; had we reached our recruitment goals and been able to complete assays during pandemic waves, then more robust methods would have been available. As more dense-data clinical trials are performed and analyzed, techniques to improve quantification of epistemological confidence will grow in tandem, much as classic biostatistics matured in the last century.

### Final thoughts

A large amount of contextual knowledge concerning the roles, associations, mechanisms, and relationships of once-obscure metabolites is becoming available (e.g., the papers we cite mentioning metabolites published in the 2020s). Many of these implicate or highlight the roles of particular metabolites in response to diet and exercise. Dense-data trials are designed to be increasingly valuable as global knowledge accumulates. Such knowledge, particularly for metabolites, is increasing rapidly; the value of dense trials (both newly conducted and re-analyses of previously released data) will also increase rapidly.

## Data Availability

Data is available in Supplemental Table 1.

## ^1^ Abbreviations

AD: Alzheimer’s disease
ADRD: Alzheimer’s disease and related disorders (or nearly equivalently: Alzheimer’s disease and related dementias)
COCOA: Coaching for Cognition in Alzheimer’s Trial
FAST: Functional Assessment Staging Test
MCI: mild cognitive impairment
MCIS: MCI Screen
RCT: randomized controlled trial
MID: minimal important difference (aka, MCID, minimal clinically important difference)
MoCA: Montreal Cognitive Assessment
MPI: Memory Performance Index

## Supplemental Table Legends

**Supplemental Table 1**. Data for all analytes. Each value is the difference between measurement at the baseline and first follow-up timepoint for an individual. Units are as described in Methods.

**Supplemental Table 2. Population-level measures for specific analytes may be causally influenced by variation in specific genes**. Many of the causal associations between genes and metabolites have been compiled (pheweb.org; Gagliano Taliun, et al., 2020). These associations relate some of the candidate mediators identified by COCOA. For example: KMO is a top influencer for both kynurenate and acetylkynurenine; SLCO1B1 is a top influencer for both hexadecanedioate and pregnenolone.

## Contributions

Planning: LH, WS, JR; Analysis: JR, LE, DM, WS; Coaching: MR; Manuscript: All authors.

## Acknowledgements

Deborah Fridman, Jennifer C. Lovejoy, Kathleen Jade, Laura Heim, Rachel Romansik, Adrienne Swietlikowski, Sheree Phillips, Maria Fischer, Dan Fischer, Lauren Dill, Michael Brant-Zawadzki, Sophiya Rajbhandari, Dale Bredesen, Mary Kay Ross, Amanda Wikan, Alex Lewis, Maia Kurnik, Christie Davidson, Jennifer Eklund, Mouna Attarha, and Mike Merzenich have inspired, managed, mooted, conceived, planned, participated, organized, supported, discussed, and/or analyzed aspects of the COCOA project or its precursors. COCOA would not have been possible without the participants, for whom we reserve the greatest acknowledgement and appreciation. Institutional support from Hoag Memorial Hospital Presbyterian was vital. Oregon Health & Science University’s Bioinformatics and Computational Biomedicine program supported interns. Data used in preparation of this article were obtained from the Alzheimer’s Disease Neuroimaging Initiative (ADNI) database (adni.loni.usc.edu). As such, the investigators within the ADNI contributed to the design and implementation of ADNI and/or provided data but did not participate in analysis or writing of this report. A complete listing of ADNI investigators can be found at adni.loni.usc.edu. Statue images (Figure 1): Folco Masi & Ziad Al Halabi (unsplash.com).

## Data Availability

Data is available in **Supplemental Table 1**.

## Supplemental Introduction: Definitions of Alzheimer’s disease (AD) and mild cognitive impairment (MCI)

The definitions of Alzheimer’s disease (AD) vary over time and between authorities. Inclusion criteria for COCOA included a Memory Performance Index (MPI) of 65 or below and a Functional Assessment Staging Test (FAST) stage between 2 and 4, inclusive (Roach, Hara, Fridman, et al., 2022), as well as an absence of a known non-AD cause of dementia. According to the Alzheimer’s Association (Alzheimer’s Association, 2022), the definitions of ‘*Alzheimer’s disease’* and ‘*Alzheimer’s dementia’* are different. AD is considered to encompass all individuals on a trajectory *towards* or within Alzheimer’s dementia; only those with actual dementia are considered to have a diagnosis of Alzheimer’s dementia. Thus, individuals with subjective or objective cognitive impairment, even with no dementia, are defined to have AD. To differentiate AD from other causes of cognitive decline, these individuals must also either have known/established biomarkers of AD or sufficient information from other sources to infer that these individuals will also have (or will eventually) known/established biomarkers of AD (e.g., amyloid or tau). Information from other sources might encompass emerging algorithms that integrate multiple assays and risk factors (e.g., Rossini et al., 2022). In some cases, one can only probabilistically assign a diagnosis of AD. Also, AD is not an exclusive diagnosis; patients may have mixed dementias. With this definition of AD, we consider the COCOA participant population to be highly enriched for AD patients. However, care must be taken in that characterization, as it can be misunderstood. For the most part, COCOA participants have AD as defined by the 2018 NIA-AA Research Framework (Jack et al., 2018), which is a similar definition to that of the Alzheimer’s Association. Although most COCOA participants were tested for AD biomarkers as part of their clinical care prior to enrollment, and all of those tested were positive, such testing was not required as part of inclusion/exclusion (due to expense). The remaining minority of participants were presumed likely to have an AD diagnosis based on an expert clinician’s judgment of their entire clinical record. It is possible that some participants in COCOA do not have AD and/or have other causes of cognitive decline — for example, some may have mixed dementias. On the spectrum of AD defined by the Alzheimer’s Association, most participants in COCOA are best described as having “very mild symptoms that may not interfere with everyday activities” and therefore are classified as “mild cognitive impairment” (MCI). However, several participants otherwise meeting the inclusion/exclusion criteria might be considered more mildly as preclinical AD or more severely as Alzheimer’s dementia. For example, someone with an MPI of 65 and a FAST of 2 could qualify for COCOA, and still be considered by clinicians to have subjective cognitive impairment (SCI) or preclinical AD. MPI scores greater than 50 are formally considered to be ‘normal’ (Shankle et al., 2009); fourteen participants had a baseline MPI score above 50 and nine never had a score less than 51 over the period of their participation. If that threshold were absolute, then scores above 50 would not be ‘objective’ measures of cognitive impairment. However, it is reasonable to consider scores of 65 and below as objective measures of impairment (for reference: an MPI score of 60 corresponds to a MoCA score of 25; some guidelines set a MoCA of 26 as the threshold for MCI). A FAST score of 2 is only subjective (not objective) functional impairment. Therefore, these individuals may be better classified with SCI than with MCI. Reasonable experts could disagree on whether COCOA individuals with a FAST of 2 and MPI score less than 65 but above 50 should be classified as SCI or MCI. Likewise, someone with dementia might still have enough function to meet all enrollment criteria (notably ability to telephonically receive coaching) and be considered by a clinician to have dementia; dementia has many facets. Seventeen participants had a baseline FAST of 4, indicative of mild dementia. In broad clinical practice, many individuals carry a diagnosis of AD without having been tested for an AD biomarker (Dubois et al., 2010). Even though COCOA inclusion/exclusion criteria may be slightly different than criteria required to meet some stringent definitions of AD, they may be well suited to recruit a real-world population of individuals diagnosed by their primary and specialist providers — which in turn implies that conclusions from COCOA may be well suited for real-world application. Knowledge from COCOA may inform not just AD, but all of ADRD.

Although the exact choice of words to describe and define the COCOA population is slightly difficult — even slightly controversial — the targeted population is currently considered by many sources to be an ideal target for interventions that slow or reverse AD decline. There are three reasons: (1) these individuals are likely to have AD — allowing efficient allocation of therapeutic resources, (2) earlier stages of pathogenesis are more likely to be interruptible or reversible than later stages (Aisen et al., 2022), and (3) individuals with early-stage AD such as those in COCOA have sufficient function to understand and respond to coaching to implement multimodal recommendations. Furthermore, solely from a perspective of trial design, to have statistical power and to complete the trial in a practical timespan, we desire to see a large and significant effect of the intervention over a short period of time. Prior to the MCI stage of the AD spectrum, control trajectories are likely to decline very slowly if at all, and a very large and/or long trial might be required. After the MCI stage, individuals may be too impaired to respond to telephonically coached multimodal interventions. If multimodal interventions can be conducted very expensively and burden free, they could theoretically be targeted at very early stages of disease, and even at entire populations. Improved early stratification could fine-tune targeting these interventions.

If our underlying and overarching hypotheses are true — that AD has multiple causes and is best treated with multiple interventions — then conclusions and clinical recommendations resulting from COCOA and similar trials are likely to be applicable to all individuals regardless of where they are on the AD spectrum. In other words, although most of our participants have MCI, we extrapolate our conclusions to all. For example, perhaps with help of caregivers, even individuals with advanced dementia are capable of modifying their diet; most are capable of exercising, pharmaceutics can be optimized, and indeed most multimodal recommendations can be adapted to the specific circumstances of each individual. Individuals at low risk may also benefit from some recommendations, such as increased exercise, which are low cost, low risk, and broadly beneficial for health.

## Supplemental Event-Driven Procedure Modifications

The text describing the primary outcome variables in clincaltrials.gov is confusing (clinicaltrials.gov/ct2/show/NCT03424200). This confusion comes from iterative editing by multiple authors to comply with cliniclatrials.gov deposition requirements; in our haste to respond to edit requests, some of these edits drew from draft rather than final trial protocols and we approved them without catching the typos. This garbled text reads:

> “Primary Outcome Measures:
>
> 1. Coaching for Cognitive Decline [Time Frame: 2 years]
>
> Test the hypothesis that coaching is better than no coaching for people in the early stages and at risk for cognitive decline. We plan to test that after two years of coaching, participants in the coaching arm will have higher cognitive scores as measured by the MCI score.
>
> Participants in COCOA will be assessed with the MCIS, which quantifies cognition with the Memory Performance Index (MPI) component score on a 0 to 100 scale (below normal: <50). The MPI will be our primary outcome measure for cognitive function. It is quick and inexpensive to administer. The MPI score is sensitive to slight cognitive deficits, and thus serves as an excellent single measure of cognition to serve as one of our two primary outcome measures Development and validation of the Memory Performance Index: reducing measurement error in recall tests.” [sic]

Although there is no second header (as might be implied by the “1” numbering in the quoted text), and we describe the MPI as our primary outcome measure, text was included in a garbled run-on sentence that indicated a second primary outcome measure. In an earlier draft protocol, FAST had also been included as a primary outcome measure before it was demoted to a secondary outcome measure in the final protocol. Elements of this draft text contributed to this mangled sentence. The strictest self-criticism would force us to conclude that we might have been able to use this garbled text for some wiggle room in a post-hoc definition of our primary outcome measure. However, the most extreme approach to addressing the criticism would be to apply a Bonferroni correction factor of 2 to the p-value for our primary outcome, converting it from p = 0.0007 to p = 0.0013. Such a correction would have no impact on any conclusion in the paper. We define the primary outcome measure clearly in Roach, Hara, Fridman, et al. (2022), but this reference was published after the trial started.

## Footnotes

2 www.medrxiv.org/content/10.1101/2022.09.27.22280385v1

