## Appendix 1. Protocol for "A multimodal intervention for Alzheimer’s disease results in multifaceted systemic effects reflected in blood and ameliorates functional and cognitive outcomes" (from another version of this preprint: version 1, 2022-09-29)

**Study Title: Coaching for Cognition in Alzheimer's (COCOA)**

PRINCIPAL INVESTIGATOR: William Shankle, M.D. (Shankle Clinic)

SUB INVESTIGATORS: Jennifer Lovejoy, Ph.D. (Institute for Systems Biology)
[redacted]
Jared Roach, M.D., Ph.D. (Institute for Systems Biology)
[redacted]

STUDY STAFF Molly Rapozo, MS, RDN, CD [redacted]

SPONSOR: Shankle Clinic

FUNDED BY: Providence St. Joseph Health Network

STUDY LOCATIONS: SHANKLE CLINIC
3900 WEST PACIFIC COAST HIGHWAY, SUITE 310
NEWPORT BEACH, CA 92663

### 1. BACKGROUND

The comprehensive treatment of cognitive impairment due to Alzheimer’s disease (**AD**) and other medical conditions requires not only pharmacologic treatment but also management of existing medical conditions and life style modifications (e.g., diet and exercise). While clinical efficacy of pharmacologic treatment has been well studied, non-pharmacologic approaches require further understanding to be clinically applied. This is partly because non-pharmacologic (i.e., lifestyle) approaches are subject to more variable compliance.

One way to address such variability is to enhance compliance with lifestyle interventions through data-driven health coaching. Health coaching may improve compliance with non-pharmacologic interventions that facilitate health, including diet, physical activity, sleep, stress management and adherence to medical treatment. It also communicates biological data that may be improved by lifestyle change, or help improve clinical management of patient medical conditions.
In this study, cognitively impaired patients will be randomly assigned to Group 1 routine care (**RC**) or Group 2 routine care plus data-driven health and wellness coaching. Each patient will be monitored for two years, and perform comprehensive evaluations to determine any differences in cognitive trajectory between the two arms. In addition, this study will evaluate “dense, dynamic data clouds (Price et al., 2017)” - consisting of clinical labs, lifestyle, genomic, proteomic, metabolomic and gut microbiome data for each study participant - using statistical and computational methods in an effort to explain the causal underpinning of any observed differences between the two arms. These

methods will be employed in an effort to explain the causal underpinning of individual trajectories, particularly if these trajectories are unusual.

The methodology of this study has been previously used in the completed 100-Person Wellness Project (**HPWP**), approved WIRB Protocol #20131979. For the HPWP, the same approach was used for data generation for a longitudinal cohort of generally healthy individuals, and were successfully able to generate biological knowledge through analysis of the dense dynamic data acquired for each individual (Price et al., 2017).

Following completion of the two-year COCOA study, all participants who consent will be followed in a 10-year longitudinal study, with less intense data gathering, to evaluate long term efficacy and residual effects of the COCOA intervention, and to enable longitudinal analysis of participant data over a longer time span.

Implementation and analyses for this study will be conducted by Providence St. Joseph Health and Shankle Clinic, the John Wayne Cancer Institute (JWCI) and JWCI's Pacific Neurosciences Institute (PNI), and the Institute for Systems Biology (**ISB**).

### 2. OBJECTIVES OF THE STUDY

One major study objective is, using 2 study arms (Group 1 RC vs Group 2 data-driven health coaching plus RC), to evaluate the efficacy of data-driven health coaching. 'RC only' will serve as the control group. Participants will be enrolled in the trial on the basis of an existence of objective cognitive impairment defined by the MCI Screen (**MCIS**) and being in one of three functional stages as defined by the Functional Assessment Staging Test (**FAST**). The three FAST stages correspond to cognitive impairment without functional impairment (FAST 2), cognitive impairment with functional impairment without impairment in instrumental activities of daily living (FAST 3, also known as mild cognitive impairment, **MCI**), or cognitive impairment with impaired instrumental activities of daily living (FAST 4). Study objectives include measuring treatment related changes in cognitive and functional abilities, quality of life, and biological or biochemical measures.

A second major study objective is to analyze longitudinal multi-omic data from individuals on a trajectory of early-stage dementia, to discover correlations between measured variables, and identify models of causation that can further advance knowledge and research in brain degeneration and healthy living.

### 3. STUDY DESIGN

#### Overview

The original study design anticipated up to 200 FAST Stages 2-4 patients with cognitive impairment to be randomized into data-driven health coaching vs. control groups. Enrollment has now stopped due to a combination of slow recruitment rate and resource limitations. Enrollment stopped after recruiting fifty three (53) participants. Nine additional individuals failed full screening, withdrew, or were removed from the study.

Participants will be treated and monitored for 24 months. **Appendix I** summarizes the study timeline.
At the end of a participant's involvement, in either group, with the primary COCOA study, they may continue to be monitored in a longitudinal study for up to 10 years. All participants, once enrolled in COCOA, are eligible to be followed longitudinally, even if they do not complete all 24 months of the primary study.

**Participant Identification and Recruitment**
Study participants have been recruited using a variety of techniques to identify participants that meet inclusion and exclusion criteria and who are able to participate in the trial, including traveling to necessary visits. Recruitment techniques include (1) identification of patients from a high-volume memory clinic (Shankle Clinic), (2) advertisement to participants in the Orange County Vital Brain Aging Program (OCVBAP), (3) establishment of a website landing page that permits interested individuals to contract the trial coordinator, and (4) radio and newsletter/newspaper advertisements.

**Procedures for Obtaining Informed Consent**
All participants will receive the Experimental Research Subject's Bill of Rights prior to signing the informed consent form (**ICF**), authorization of use and disclosure of protected health information (**PHI**), and authorization of medical record release for the participant's treating physician, will be obtained from each participant prior to enrolling in the study. A copy of all signed ICF's will be given to the participant, and the investigator will retain the original.

FAST staging (see **Attachment II** for the FAST administrating and scoring) will be done before participants are consented to determine whether they or a legally appointed representative (**LAR**) can consent to participate in the study. In this study, participants with FAST stages 2-4 will be recruited (see Inclusion Criteria).

- <u>Considerations for consenting FAST stages 2-3 participants:</u> FAST stages 2-3 patients are usually competent to make medical and legal decisions, and will be consented directly, unless there is a caregiver, legally appointed representative, or other reason to think that an informed consent cannot be given by the patient without approval by a reliable informant acting on their behalf.

- <u>Considerations for consenting FAST stage 4 (mild dementia) participants:</u> FAST stage 4 patients will be consented by having them give oral or written *assent*, indicating their preference with regard to study participation. In addition, the caregiver or legally appointed representative of a demented patient will be consented to assure full understanding of study procedures and willingness on behalf of the patient to participate in the study.

This approach has been evaluated by the Department of Psychiatry and Behavioral Sciences at Johns Hopkins, and found to adequately ensure informed consent (Black et al. 2010). They concluded that ADRD patients should not be excluded from study participation if they cannot directly consent themselves, so long as their caregivers can consent, and the patient can assent, either verbally or in writing, their preference to participate in the study.

The caregiver or legally appointed representative will be required to accompany participants to participate in the required procedures. A copy of the appropriate document (e.g., the power of attorney for healthcare) will be obtained and filed with the original ICF.

**Screening and Enrollment**
Participants interested in participating in the study will be screened based on the inclusion and exclusion criteria listed below.

<u>Inclusion Criteria</u>

- Participants must be age 50 and older (no age upper limit) at time of informed consent.
- Adults of any gender, race or ethnicity are eligible to enroll in the study.
- Participants must have cognitive impairment as demonstrated by a Memory Performance Index (MPI) of 65 or below.
- Participants must have a FAST stage of 2-4.
- Participants with FAST Stage 4 must have the caregiver or legally appointed representative who can provide an appropriate documents (e.g., the power of attorney for healthcare)
- Participants must be proficient in spoken and written English for consenting as well as for study participation since the intervention in this study (e.g., coaching program) is currently only available in English.
- Participants with medical conditions must be stable for these conditions. Stable control on medication is acceptable.
- Participants, with or without assistance, must be able to use a computer and web interface. If assistance is needed, it must be readily available to them.
- Participants must be able to converse with a coach telephonically. Telephone coaching is part of data-driven health coaching.
- Participants must have regular access to a computer and the Internet along with dedicated email address since certain aspects of the program (e.g. cognitive training) are delivered electronically.
- Participants must have normal visual acuity (or corrected to normal) and normal color vision as indicated by self-report
- Participants must have adequate hearing acuity as indicated by self-report
- Participants must have adequate motor capacity to use a mobile device/iPad/computer as indicated by self-report

Exclusion Criteria

- Participants with an existing diagnosis of a non-AD neurodegenerative disorder (e.g., Lewy Body Disease, Frontal-Temporal Disease).
- Participants with a diagnosis of cerebrovascular disease as the primary cause of cognitive impairment.
- Participants who have participated in the Arivale program or the HPWP. These employed coaching similar to the approach that will be used in the current study.
- A previously reported AD high-risk mutation (e.g., in the PSEN or APP genes) in the participant or immediate family (children, siblings, or parents). Such patients may accumulate amyloid faster than in late onset AD, and therefore may show less pronounced benefit from intervention.

**Randomization**
A computerized randomized block design will be used for participants who meet inclusion and exclusion criteria. The block randomization is designed to enroll half the participants in Group 1 and half in Group 2.

All enrolled participants will undergo:
- Biological data collection
- Life style and quality of life data collection
- Medical record collection
- Cognitive assessment
- Functional assessment
- Neurological and physical evaluation

In addition to the above data collection and assessments, Group 2 will receive data-driven health coaching as described below.

**Data-Driven Health Coaching**
Data-driven health coaching is provided to participants in Group 2. Currently this coach is based at PSJH's PNI affiliate. The coach will facilitate interpretation and communication of participants' genomic and other biological data. Although changing coaching staff is not expected during this trial, all current and future coaches will be currently licensed allied health providers including registered dietitians or registered nurses. Coaching calls typically occur on monthly basis, with email and text communication occurring between the monthly calls (as often as weekly depending on participant preference and responsiveness). If clinically necessary and by mutual agreement with the participant, coaching calls and additional health assessments may occur more or less frequently. There is no face-to-face coaching provided. Participants may contact their coach at any time with questions or for support. Using the combination of an individuals' biological data (including genomics, blood, stool, saliva etc.), activity data, behavioral data and other incoming streams of health information the coach will support adoption and maintenance of healthy behaviors including healthy nutrition, physical activity, stress management, weight management, and adherence to doctor-prescribed medical regimens. All

information to support healthy behavior will be based on standard, published, and well-researched health data. Also, non-urgent out-of-range labs will be discussed on the monthly phone call with the coach, or via other regular communication. Coaches will not give any medical advice, unless licensed to do so, and will refer participants to their primary healthcare provider for medical conditions discovered from the data. Because all participants will be managing cognitive decline (an enrollment criterion for the study), coaching will include recommendations for exercise, diet, and brain training for all individuals. Brain training for this study will be implemented with Posit's BrainHQ web-based training tools (brainhq.com), or other appropriate brain-training (e.g., Dakim). Coaches will have access to participants BrainHQ or other cognitive training data to facilitate coaching (e.g. to encourage compliance with the recommend training regimen).

**Participant ID Assignment**
Upon enrollment, each patient will be assigned a study ID that will be used to link all data. Only the PI/Sub-PI, and designated members of the research team will have access to a method to link each patient with their participant ID. This spreadsheet will be password protected and reside within the institution's HIPAA compliant system.

**Fees for Participation**
There are no costs associated with participating in this research. Routine medical care will be billed to the participant's insurance.

**Compensation**
All participants will be compensated $25.00 per completed office visit and $35.00 per completed laboratory visits to cover time, travel and inconvenience. If the participant completes all study visits and collections, the maximum amount they will receive is $385.

Additionally, upon completion of the primary study, Group 1 participants will receive 3 coaching calls plus email/text coaching at the end of the trial.

Participants may continue to be followed longitudinally after their involvement primary COCOA study. Such participants may be invited to one office visit and one associated blood draw per year for up to 10 years. They could thus receive up to $600 in additional compensation if they complete all of these visits.

### 5. EVALUATIONS AND ASSESSMENTS

#### Biological Data Collection

<u>Genetic Data</u>
At baseline (one time only), high quality genomic DNA will be extracted from blood and shipped to an independent, CLIA-certified genomics vendor. Genotyping and/or whole-genome sequence will be acquired from each individual. Genomic results are then returned to PSJH. Any data with HIPAA identifiers will be maintained with the PSJH system. Data will be analyzed and interpreted at PSJH, JWCI, and ISB. A subset of CLIA-certified gene variants actionable through nutritional or lifestyle intervention will be reported to participants. These include genetic variants related to Health Dimensions such

as: Heart Health (e.g. atherosclerosis, hypertension), Diabetes Risk (insulin resistance), Inflammation, Nutrition, Health Aging (oxidative stress and toxins) and Stress. Representative genes currently reported include PPARG (diabetes risk), AGT (salt sensitivity and blood pressure), and GSTT1 (glutathione metabolism). Additionally, "polygenic profiles" will be reported based on hundreds of genes that have been previously studied and shown to relate to predisposition for complex phenotypes like body weight, injury risk during exercise, or blood lipids. However, since understanding of the significance of genomic variants is dynamic, specific polymorphisms and profiles reported to participants are subject to modification if literature supports refinement of the genetic report.

Clinical Chemistry Data

Blood will be collected and assayed at baseline and at 4, 8, 12, 18 and 24 months at Shankle Clinic or any of a number of phlebotomy centers run by a CLIA-certified laboratory (LabCorp, Inc.). Licensed phlebotomists will draw the blood. The maximum blood to be collected at any one time from participants is 150 mL (about 10 tablespoons). By comparison, the amount of blood collected when people donate blood is 1 pint, or roughly 500 mL. Chemistries assayed may include analyses of any blood component including proteins, metabolites, nucleic acids and contents of exosomes.

**Participants will receive the following instructions;**

**4 days in advance:**

- Unlike your first blood draw, please continue to use your nutritional supplements prior to this draw.
- **Discontinue** non-prescription medications, including acetaminophen (Tylenol), ibuprofen (Advil) and over-the-counter cold remedies. Please continue taking low-dose aspirin if you do so for your heart health.

  *If you have any concerns about discontinuing non-prescription medications, please consult with your physician prior to discontinuing.*
- **Do not discontinue** any prescription medications or nutritional supplements.

**24 hours in advance:**

- Do not drink alcohol.
- Avoid vigorous exercise.
- Do not consume products containing aspartame (e.g. Nutra-Sweet, Equal, various sugar-free products) or monosodium glutamate (MSG), a flavor enhancer found in many processed and restaurant foods.

**12 hours in advance:**

- **Begin fasting 12 hours prior to your blood draw.**
- No food or drink (except water). This includes chewing gum, mints, cough drops, black coffee, and tea.

- **Drink plenty of water.** Hydrated blood vessels facilitate a quick and comfortable blood draw.

Stool and Saliva Samples

Stool samples for gut microbiome composition and saliva samples for stress hormone levels will be collected at baseline and at 12 and 24 months by the participants at home using a specimen collection pack provided by third-party vendors. Samples will be shipped directly to independent CLIA-certified lab vendors. Collection packages will contain pre-printed coded identification and shipping labels. Participants who fail to return bio specimen collection will be sent reminders until completed. Saliva collection will occur four (4) times on a single day, with the first sample before 7:00 a.m., and within an hour of waking, and last sample between 9-10 p.m.

**Life Style and Quality of Life Data Collection**

Lifestyle Data

Physical activity, heart rate, and sleep data will be collected automatically and on a nearly continuous basis by using a FitBit tracker, or similar commercially-available tracker, which will be provided to each participant in the study.

Anthropometrics

Body weight and waist circumference will be collected at baseline and at baseline and at 4, 8, 12, 18 and 24 months at the time of the blood draw either by clinical lab personnel or by the study participant. Coaches may recommend that participants also measure weight more frequently at home (e.g. if they are working on weight loss).

Questionnaires (see **Attachment 1**)

Questionnaire Data will be collected from the participants electronically at baseline, and subsequently at varying intervals. Questionnaires are completed online at baseline and at 4, 8, 12, 18 and 24 months except as indicated below. Participants can opt out of these at any time if they choose.

1. The "Big Five" Personality Survey (Baseline only). A validated personality questionnaire assessing five dimensions of personality. Personality type from this questionnaire has been shown in other studies to be a metric of health or illness.
2. Perceived Stress Scale (PSS-4). A validated measure of perceived stress.
3. Oxford Happiness Scale. A validated measure of an individual's degree of happiness and life satisfaction. Research in positive psychology shows that traits such as happiness are linked to health outcomes.
4. Personal and Family Medical History and Vitals (Baseline, 12 and 24 months). This information is used both for the tailored coaching and as a factor in analyzing genetic results. Vitals include height, weight, medication and supplement usage and coaching preferences.
5. Digestion Questionnaire. A brief questionnaire assessing digestive symptoms used in coaching.

6. Lifestyle Questionnaire. A questionnaire assessing various aspects of lifestyle including diet, exercise, sleep, alcohol and drug use and social and environmental factors.
7. Journey Assessment. A questionnaire that asks participants about their wellness goals from participating in the coaching program and current attitudes about their own wellness and degree of satisfaction with their health.
8. Satisfaction Survey. A survey to assess participation satisfaction with the coaching services they are receiving.
9. Depression Questionnaire. Patient Health Questionnaire-9 (PHQ-9).

**Cognitive Training**

Data from BrainHQ will be collected, including the frequency and duration of training for each training exercise, and the score for each exercise. If the participant is pursuing an alternative cognitive training program, similar data will be collected, if it is available.

**Medical Record Collection**

A patient’s medical record will be collected from their treating physician office (i.e., for cognition or neurological issues, possibly their primary care provider) at the screening/baseline, 12 month, and 24 month visits for medication type, dosage and frequencies as well as other existing medical conditions and their treatment. The data will be entered into an electronic data capture system by research staff at Shankle Clinic. In the event that a particular participant’s data has extreme or outlier characteristics, their medical record will be reviewed to identify explanatory factors.

**Cognitive Assessments**

Each participant will be assessed for cognitive and functional abilities using the:

- MCI Screen (MCIS). Administered at baseline, 4, 8, 12, 18 and 24 months.
- Healthy Brain Checklist. Administered at baseline, 4, 8, 12, 18 and 24 months.
- The Delis-Kaplan Executive Functioning System (D-KEFS). Administered at baseline, 12 and 24 months.
- The Montreal Cognitive Assessment (MoCA). Administered at baseline, 12 and 24 months with 2 weeks delay from other cognitive assessments to avoid interference effects.
- BrainHQ assessment.

The PI and/or test administrator may choose not to administer the D-KEFS, or to administer a shorter version of the D-KEFS, if a participant is felt to be unable to effectively complete the full battery or tests, or if completing the full battery would place an undue burden (e.g., due to anxiety or limited stamina) on the participant.

See **Appendix II** for more details on cognitive assessment tools.

**Functional Assessment**

The Functional Assessment Staging Test (**FAST**) will be used to assess functional abilities at baseline and at 12 and 24 months. The FAST is an internationally validated, semiquantitative measure of the functional changes in AD from the entirely asymptomatic stage to severe dementia. There are 7 major stages and 16 sub-stages, each with a published mean duration of the untreated course of AD. See **Appendix III** for more details on the FAST.

**Neurological and Physical Evaluation**
Neurological and physical evaluation will be conducted at baseline to document:

- Any abnormal physical or neurological signs
- Whether they are considered to be clinically significant

### 6. LONGITUDINAL FOLLOWUP STUDY

Following participation in the primary COCOA study (typically 24 months from time of enrollment), participants may continue to participate in a longitudinal observational study of past COCOA participants. There will be no coaching intervention during this longitudinal study, although participants may continue or initiate any of the recommendations or interventions they may have received or learned during the COCOA study (e.g., exercise, diet, cognitive training). BrainHQ for all participants (including Group 1) will continue to be made available. Medical records will be collected for any cognitive test results. Participants may be invited in the follow up study for an office visit once per year for cognitive testing and a blood draw. A subset of the tests will be assayed and used during the primary COCOA Study. The DK-EFS will not be administered. Stool or saliva will not be collected.

### 7. DATA AND SAMPLE STORAGE AND MONITORING

#### Data Collection and Storage

Biological samples may be stored for up to 50 years at a PSJH biostorage facility. A code number will be assigned to all samples. Participants' name, medical record number, or other identifying information will not be stored with the samples. Participants' names and contact information will be stored at Shankle Clinic and PNI and not be shared with researchers outside of the PSJH system. Researchers, other than those providing direct medical care as part of RC, will not have access to personally identifying information.

For all biological and questionnaire data described above, but not medical records, they will be collected into a centralized secure database and protected by technological, physical and administrative safeguards as provided by HIPAA and other applicable privacy laws. Only PSJH coaches, clinical staff and authorized operations personnel will have access to this data. Data may also be de-identified, then stored in a separate secure database for potential future use. Biological specimens such as plasma and purified DNA will be placed in biostorage to enable future research. Samples will be stored at JWCI, ISB, and/or a commercial vendor (currently Brooks Biostorage). All samples will be stored as coded samples with no personal identifiers. Codes will be stored separately, password protected and only accessible to authorized personnel.

All cognitive assessment data and medical information collected from participant medical records (e.g., medication and existing medical conditions) will be entered in an electronic data capture system. Research data and de-identified medical information may be stored for up to 50 years.

**Data Analysis and Data Monitoring**

The process of analyzing participant study data may continue indefinitely. Analysis of the incoming participant data using multiple regression analysis for statistical changes in clinical health markers over time. Discovery research on interactions between data sets will rely on standard methods for analysis of big data sets. Only individuals who have completed all applicable HIPAA training will be provided access to PHI.

**Data Storage and Confidentiality**

All data will be kept private and shared with the participant as outlined above. Participants' biological data is stored in one database with one set of security credentials and Personally Identifiable Information is stored on a different database with separate security credentials. All data will be stored in a password-protected secured server and only specifically identified study personnel will have access to the data. Discoveries derived from participant data that may be published and shared publicly will have all data de-identified.

### 8. ANALYTICAL METHODS

In order to determine whether treatment arms have differing rates of cognitive decline over time, linear mixed-effect models with random intercepts and slopes will be performed, such that individual patients could both 1) have differing starting trajectories, or intercepts; and b) have differing rates of cognitive decline over time, or slopes. The estimated mean difference between treatment arms in rate of cognitive decline (measured via MPI) will be assessed by including an interaction term between time and treatment arm. Covariates (fixed effects) will include: gender, race, and education; and baseline indicators for age, physical activity, and presence of vascular comorbidities, depression history, self-rated health, and MPI. Generalized estimating equations (GEE) will be performed with an exchangeable correlation matrix, including the same covariates and interaction terms, as confirmatory analyses. Hierarchical Bayesian Cognitive Processing Models will be employed to measure underlying cognitive processing parameters.

Our experience with the HPWP, it was observed that many new relationships that were statistically significant from a population of this size. Each patient has multiple time points, which enhances our statistical power as well. Therefore, identification of associations between clinical outcomes (i.e., MPI) and molecular variables is anticipated. ISB has extensive expertise in a wide range of statistical learning approaches that will be applied to these data to identify novel relationships that can be further verified via subsequent experiments and trials to follow this one.

Exploratory integrative data analysis will be performed. These analyses may gain much deeper insights into the biology underlying clinical processes. Multiple machine learning techniques will be applied to calculate disease risk scores and develop predictive classification models (e.g., sparse logic models, sparse linear models, relative expression reversal algorithms that have been pioneered). The robustness of such models will be rigorously tested by test set holdouts, cross-validation, feature dropout, predicting data of held-out future time points based on observed trends, and gauging the robustness of models over time. Novel approaches are being developed for studying sets of atypical values (e.g. outliers) that appear in individuals as a means for identifying perturbed biological networks. Functional analysis of these outliers can – even on an individual basis – point to elements of a patient's biology that have been pushed into abnormal states, a potential sign of pathologies.

### 9. PROCEDURES FOR STUDY DOCUMENTATION

#### Informed Consent

A copy of all signed informed consent form (ICF's) will be given to the participant, and the investigator will retain the originals. **Section 3** provides further details regarding the specific requirements for informed consent.

#### Investigator Study File

Documentation concerning the study including, but not limited to, study protocol and any amendments, the IRB documents, study logs, monitoring activities and correspondences, case report forms (CRFs) will be kept on-site in secure location.

#### Monitoring and Quality Assurance

Quality assurance procedures will include training of all study personnel, monitoring of data and reviews to verify that all elements of Good Clinical Practice (GCP) guidelines and FDA regulations are met. Monitoring will be conducted to assure that data are accurate and in agreement with source documentation; verify that consent for study has been properly obtained and documented; confirm that participants enrolled in the study meet inclusion and exclusion criteria; monitor for safety (e.g. adverse events) and assure that all essential documentation required by GCP guidelines are appropriately filed.

#### Retention of Study Documents

Study documents will be retained for a period of two years following completion of the study. No study documents will be destroyed or moved to a new location without prior written approval from the sponsor. If the investigator relocates, retires, or withdraws from the clinical study for any reason, all records required to be maintained for the study will be transferred to an agreed-upon designee, such as another investigator at the institution where the study was conducted. Medical or coaching records used for treatment purposes will be retained separately as required by state data retention laws, HIPAA and other applicable laws.

### 10. PROCEDURES FOR HANDLING ADVERSE EVENTS (AEs) AND SERIOUS ADVERSE EVENTS (SAEs)

Due to the nature of this study, adverse events are not expected of the same nature as might be expected if an investigational pharmaceutical were being investigated. Interventions in this study include routine care and coaching, neither one of which is likely to include a physiological adverse event. However, adverse events could still occur to participants, including adverse events not related to the study. Suicide or attempted suicide would be an example of an adverse event.

**Definition of an AE**

The following definition of an adverse event (**AE**) will be used for this study:

- Any untoward medical occurrence in an enrolled participant who underwent procedures required by the study.

An AE can be any unfavorable and unintended sign, symptom, or disease (new or exacerbated) occurring after starting the procedures required by the study, even if the event is considered not related to the study procedures.

As far as possible, each adverse event will also be described by its duration (start and end dates), the intensity, its expectedness related to the known risks associated with the study procedures, and the action(s) taken.

It is the responsibility of the investigator to perform periodic and special assessments for AEs. The investigator and clinical staff will note all AEs reported by the participants. All clinical complaints volunteered by, or elicited from, the participant during the study will be recorded separately on the appropriate page of the CRF. If any AE occurs, the participant will receive appropriate treatment and medical supervision if warranted.

All AEs judged to be clinically significant will be followed until resolution.

**Definition of an SAE**

The definition of a serious adverse event (**SAE**) in this study is an AE that meets any of the following criteria:

- Results in death
- Is life-threatening

NOTE: The term 'life-threatening' in the definition of 'serious' refers to an event in which the participant was at risk of death at the time of the event. It does not refer to an event, which hypothetically might have caused death if it were more severe.

- Requires hospitalization or a prolongation of an existing hospitalization

NOTE: In general, hospitalization signifies that the participant has been detained at the hospital or emergency ward for observation or treatment that would not have been appropriate in the physician's office or outpatient setting. Complications that occur during hospitalization are AEs but not necessarily SAEs. An occurrence or complication that prolongs hospitalization is an SAE. When in doubt as to whether hospitalization occurred or was necessary, the AE should be considered an SAE. Hospitalization for elective treatments of a pre-existing condition that did not worsen from its original baseline is not considered an SAE.

- A persistent or significant disability or incapacity

NOTE: The term disability means a substantial disruption of a person's ability to conduct his/her regular life functions. This definition is not intended to include AEs of relatively minor medical significance such as uncomplicated headache, nausea, vomiting, diarrhea, influenza and accidental trauma (e.g., sprained ankle) which may interfere or prevent everyday life functions but do not constitute a substantial disruption.

- Other important medical event

NOTE: Medical or scientific judgment should be exercised in deciding whether reporting is appropriate for other important medical events that may not result in death, be life-threatening, or require hospitalization but still may jeopardize the participant or may require medical intervention to prevent one of the outcomes listed in this definition. These events should also be considered serious.

**Recording AEs and SAEs**

If an AE or SAE occurs, and it is deemed related and unanticipated to this study, it is the responsibility of the PI to review all documentation (e.g., hospital progress notes, laboratory, and diagnostic reports) relative to the event(s). The PI is to record all relevant information regarding any AE (including SAEs) on the AE page of the CRF. The PI will also attempt to establish a diagnosis of the event based on the signs, symptoms, or other clinical information. In such cases, the diagnosis, not the individual signs and symptoms, should be documented on the appropriate CRF as the AE or SAE.

**Assessment of Intensity**

The PI will make an assessment of intensity for each AE and SAE reported during the study. The assessment will be based on the investigator's clinical judgment. The intensity of each AE recorded in the CRF should be assigned to one of the categories shown in **Table 2**.

**Table 2**: Classification of AEs by Intensity

| Intensity | Definition |
|---|---|
| Mild AE | An event that is easily tolerated by the participant, causing minimal discomfort and not interfering with everyday activities. |
| Moderate AE | An event that is sufficiently discomforting, causing interference with normal everyday activities. |
| Severe AE | An event that prevents the participant from performing their everyday normal activities. |

Any AE that changes in intensity or grade during its course will be recorded on the CRF at the highest-level experienced by the participant.

NOTE: An AE that is assessed as severe should not be confused with an SAE. Severity is a category utilized for rating the intensity of an event (such as mild, moderate, or severe myocardial infarction. However, the event itself may be of relatively minor medical significance, such as a severe headache), and both AE(s) and SAE(s) can be assessed as severe.

**Expectedness of AEs**
An expected AE is one that is consistent with the known risk information associated with the study procedures.

An unexpected AE is defined, as any AE, the specificity or severity of which is not consistent with the known risk information. "Unexpected," as used in this definition, refers to an AE that has not been previously observed rather than from the perspective of such an AE not being anticipated based on the known effects of procedures required by the study.

**Follow-Up of AEs and SAEs**
After the AE or SAE report, the PI will proactively follow each participant.

All AE(s) and SAE(s) will be followed until:

- Resolution
- The condition stabilizes
- The event is otherwise explained
- The participant is lost to follow-up

Once the event is resolved, the appropriate SAE report page will be updated. The PI will also ensure that the follow-up includes any supplemental information that may explain the causality of the event(s).

### 11. CONFIDENTIALITY

#### Data
All information regarding the nature of the study provided by the sponsor or study monitor to the investigator (with the exception of information required by law or regulations to be disclosed to the IRB or the participant) must be kept in confidence by the investigator.

#### Participant Anonymity
The anonymity of the participants must be maintained. Participants will be identified by their initials and an assigned participant number on CRFs and other documents retrieved from the site or sent to the study monitor, sponsor, regulatory agencies or central laboratories/reviewers. Documents that identify the participant (e.g., the signed informed

consent document) must be maintained in strict confidence by the investigator, except to the extent necessary to allow auditing by the appropriate authorities, the study monitor, or sponsor representatives.

### 12. PARTICIPANT DISCONTINUATION AND TERMINATION OF STUDY

Although participants will be encouraged to complete the study, they may voluntarily withdraw at any time. The investigator will document the reasons for discontinuation.

Also the investigator may withdraw the participant from the study. The participant will be removed from the study without their consent for various health, safety or administrative reasons, including but not limited to the following:

- The investigator decides that continuing in the study would be harmful to participant.
- The sponsor cancels the study.
- Other administrative reasons.

The investigator will make the decision and let the participant know if it is not possible for him/her to participate.

### APPENDIX I: STUDY TIMELINE (IN MONTH)

| | Screen. | Randmz. | Baseline | 2 | 3 | 4 | 5 | 6 | 7 | 8 | 9 | 10 | 11 | 12 | 13 | 14 | 15 | 16 | 17 | 18 | 19 | 20 | 21 | 22 | 23 | 24 |
|---|---|---|---|---|---|---|---|---|---|---|---|---|---|---|---|---|---|---|---|---|---|---|---|---|---|---|
| **Enrollment / Randomization** | | | | | | | | | | | | | | | | | | | | | | | | | | |
| Consent & Screening | x | | | | | | | | | | | | | | | | | | | | | | | | | |
| Randomization | | x | | | | | | | | | | | | | | | | | | | | | | | | |
| **Intervention** | | | | | | | | | | | | | | | | | | | | | | | | | | |
| Wellness Coaching | | | x | x | x | x | x | x | x | x | x | x | x | x | x | x | x | x | x | x | x | x | x | x | x | x |
| **Clinical Data Collection** | | | | | | | | | | | | | | | | | | | | | | | | | | |
| Medical Record Release/Collection | x | | | | | | | | | | | | | x | | | | | | | | | | | | x |
| **Neuro/Physical Evaluation** | | | | | | | | | | | | | | | | | | | | | | | | | | |
| Standard neuro/physical evaluation | | | x | | | | | | | | | | | | | | | | | | | | | | | |
| **Biological Sample Collection** | | | | | | | | | | | | | | | | | | | | | | | | | | |
| Clinical Chemistry Data | | | x | | | x | | | | x | | | | x | | | | | | x | | | | | | x |
| Stool and Saliva Samples | | | x | | | | | | | | | | | x | | | | | | | | | | | | x |
| Lifestyle Data | | | x | x | x | x | x | x | x | x | x | x | x | x | x | x | x | x | x | x | x | x | x | x | x | x |
| Anthropometrics | | | x | | | x | | | | x | | | | x | | | | | | x | | | | | | x |
| Participant Questionnaires | | | x | | | x | | | | x | | | | x | | | | | | x | | | | | | x |
| **Neuropsychologic Assessments** | | | | | | | | | | | | | | | | | | | | | | | | | | |
| MCI Screen | x | | x | | | x | | | | x | | | | x | | | | | | x | | | | | | x |
| Healthy Brain Checklist | | | x | | | x | | | | x | | | | x | | | | | | x | | | | | | x |
| The Delis-Kaplan Executive Functioning System | | | x | | | | | | | | | | | x | | | | | | | | | | | | x |
| MOCA | | | x | | | | | | | | | | | x | | | | | | | | | | | | x |
| **Functional Assessments** | | | | | | | | | | | | | | | | | | | | | | | | | | |
| Functional Staging Assessment Test (FAST) | x | | x | | | | | | | | | | | x | | | | | | | | | | | | x |

### APPENDIX II: COGNITIVE ASSESSMENTS

#### MCI Screen

The MCI Screen (**MCIS**) was partly derived from the 10-word recall test of the CERAD battery or ADAS-Cog, but has been substantively enhanced to measure more precisely the following tasks and cognitive domains.

- Context-Free Immediate Recall Task, which measures:
    - Attention and working memory (Left dorsolateral prefrontal cortex).
    - Encoding (entorhinal-perirhinal and posterior cingulate cortex).
- Context-Free Delayed Recall and Recognition Task, which measures:
    - Retrieval (entorhinal-perirhinal and posterior cingulate cortex).
    - Encoding (entorhinal-perirhinal and posterior cingulate cortex).
- Context-Rich Delayed Recall Task of the 9 Animals from the Triadic Comparisons Task, which measures retrieval (hippocampus and posterior cingulate cortex).
- Prediction of Context-Free Delayed Recall Performance, which measures judgment and awareness of memory ability (Metamemory: ventromedial prefrontal cortex).
- Triadic Comparisons of Animals Task, which measures judgment of semantic differences from a single category of items (Left Inferior Prefrontal Cortex, at least).

The MCIS is enhanced with an advanced scoring method for higher accuracy (**Table A**), with 15 additional wordlists that are linguistically equivalent and measure context-free recall by minimizing inter-item associability within each list. The MCIS is also enhanced by incorporating measures of semantic judgment, of metamemory, and of context-rich delayed free recall. The MCIS measures a wide range of cognitive impairment, from normal to moderately severe dementia, and is administered with high reliability by office staff after one or two practice sessions using a computer-guided interface for consistent test administration.

**Table A: MCI Screen Accuracy by Severity**

| | Overall Accuracy | Sensitivity | Specificity |
|---|---|---|---|
| MCI vs. Normal | 97% | 95% | 88% |
| MCI/Mild Dementia vs. Normal | 98% | 97% | 88% |
| Mild Dementia vs. Normal | 99% | 96% | 99% |

The MCIS generates the Memory Performance Index (**MPI**) that quantifies a patient's pattern of context-free recall during learning and after a delay, on a scale ranging from 0-100. The MPI classifies the score as either Normal or Below Normal according to the participant's demographic peer group.

**The Delis-Kaplan Executive Functioning System**

The Delis-Kaplan Executive Function System (D-KEFS; Delis, Kaplan, & Kramer, 2001) is specifically designed for the assessment of executive functions including flexibility of thinking, inhibition, problem solving, planning, impulse control, concept formation, abstract thinking, and creativity. The D-KEFS, composed of nine standalone tests that can be individually or group administered, provides a standardized assessment of executive functions in children and adults between the ages of 8 and 89.

**The Montreal Cognitive Assessment (MoCA)**

The MoCA is a 10-15 minute assessment, similar to the Mini-Mental State Exam, consists a few brief tasks measuring several cognitive domains. The domains measured include constructional praxis, set-shifting, object naming, attention, immediate and delayed free recall, orientation to time and place, sentence repetition, category fluency, plus simple analogies.

**Healthy Brain Checklist (HBC)**

The HBC is a 16-item, subjective assessment of changes in memory that are sensitive to either normal aging or mild cognitive impairment. The HBC was derived from a review of the literature for questions with high factor loading for either age-related memory changes or mild cognitive impairment. The HBC has been psychometrically validated using item response theory. In contrast to objective testing, which measures cognitive performance at a single time point, the HBC measures change in memory over time. In this regard, the HBC is complementary to the MPI, such that the combination of both subjective and objective memory results characterizes memory severity and change more precisely than either measure alone.

**BrainHQ Assessments**

Online assessments of brain speed, people skills, navigation, attention, intelligence, and memory created by Posit (www.brainhq.com) will be self-administered through a web interface every 6-12 months. It will take about half an hour to complete these assessments.

### APPENDIX III: THE FUNCTIONAL ASSESSMENT STAGING TOOL

The Functional Assessment Staging (**FAST**) procedure is an internationally validated measure of the progression of Alzheimer's disease, from the completely asymptomatic stage to severe dementia. The FAST staging procedure also accurately characterizes the progression of other dementing disorders, including vascular dementia and other non-AD disorders. It classifies functional ability into 7 major stages, with a total of 16 sub-stages. Each FAST stage is associated with a mean duration, in months, for untreated AD patients. These mean durations are useful for normalizing the duration that a given patient spends in a given FAST stage. The mean duration of a FAST stage can also be used to help determine whether any given treatment of a patient may be delaying their disease progression. Each FAST stage is also associated with an MMSE range of scores and a developmental age range (emotional and reasoning age) as a reference.

| Stage | Severity | Characteristic | Expected Untreated AD Duration (months) | Mental Age (years) | MMSE |
|---|---|---|---|---|---|
| 1 | Normal Aging | No deficits whatsoever | - | Adult | 29-30 |
| 2 | Subjective Cognitive Impairment (SCI) | Subjective functional deficit | - | Adult | 28-29 |
| 3 | Mild Cognitive Impairment (MCI) | Objective functional deficit interferes with a person's most complex tasks | 84 | 12+ | 24-28 |
| 4 | Mild Dementia | IADLs become affected, such as bill paying, cooking, cleaning, traveling | 24 | 8-12 | 19-20 |
| 5 | Moderate Dementia | Needs help selecting proper attire | 18 | 5-7 | 15 |
| 6a | Moderately Severe Dementia | Needs help putting on clothes | 4.8 | 5 | 9 |
| 6b | Moderately Severe Dementia | Needs help bathing | 4.8 | 4 | 8 |
| 6c | Moderately Severe Dementia | Needs help toileting | 4.8 | 4 | 5 |
| 6d | Moderately Severe Dementia | Urinary incontinence | 3.6 | 3-4 | 3 |
| 6e | Moderately Severe Dementia | Fecal incontinence | 9.6 | 2-3 | 1 |
| 7a | Severe Dementia | Speaks 5-6 words during day | 12 | 1.25 | 0 |
| 7b | Severe Dementia | Speaks only 1 word clearly | 18 | 1 | 0 |

| | | | | | |
|---|---|---|---|---|---|
| 7c | Severe Dementia | Can no longer walk | 12 | 1 | 0 |
| 7d | Severe Dementia | Can no longer sit up | 12 | 0.5-0.8 | 0 |
| 7e | Severe Dementia | Can no longer smile | 18 | 0.2-0.4 | 0 |
| 7f | Severe Dementia | Can no longer hold up head | 12+ | 0-0.2 | 0 |
