## Appendix 2. CONSORT Checklist for "A multimodal intervention for Alzheimer’s disease results in multifaceted systemic effects reflected in blood and ameliorates functional and cognitive outcomes" (from another version of this preprint: version 1, 2022-09-29)

### CONSORT 2010 checklist of information to include when reporting a randomised trial

| Section/Topic | Item No | Checklist item | Reported on page No |
|---|---|---|---|
| **Title and abstract** | | | |
| | 1a | Identification as a randomised trial in the title | Not applicable to follow-up systems analysis paper and/or would make for an overly long title. |
| | 1b | Structured summary of trial design, methods, results, and conclusions (for specific guidance see CONSORT for abstracts) | Not applicable to follow-up systems analysis paper, although provided in streamlined form to limit the need to consult the previous publication. |
| **Introduction** | | | |
| Background and objectives | 2a | Scientific background and explanation of rationale | Introduction |
| | 2b | Specific objectives or hypotheses | Introduction |
| **Methods** | | | |
| Trial design | 3a | Description of trial design (such as parallel, factorial) including allocation ratio | See previously published Methods Paper (Roach, Hara, Fridman, et al., 2022). |
| | 3b | Important changes to methods after trial commencement (such as eligibility criteria), with reasons | Event Driven Procedural Modifications |
| Participants | 4a | Eligibility criteria for participants | See previously published Methods Paper. |
| | 4b | Settings and locations where the data were collected | See previously published Methods Paper. |
| Interventions | 5 | The interventions for each group with sufficient details to allow replication, including how and when they were actually administered | See previously published Methods Paper. |
| Outcomes | 6a | Completely defined pre-specified primary and secondary outcome measures, including how and when they were assessed | See previously published Methods Paper. |
| | 6b | Any changes to trial outcomes after the trial commenced, with reasons | See Event Driven Procedural Modifications |
| Sample size | 7a | How sample size was determined | See previously published Methods Paper. |
| | 7b | When applicable, explanation of any interim analyses and stopping guidelines | N/A |
| Randomisation: | | | |
| Sequence generation | 8a | Method used to generate the random allocation sequence | See previously published Methods Paper. |
| | 8b | Type of randomisation; details of any restriction (such as blocking and block size) | See previously published Methods Paper. |
| Allocation concealment mechanism | 9 | Mechanism used to implement the random allocation sequence (such as sequentially numbered containers), describing any steps taken to conceal the sequence until interventions were assigned | See previously published Methods Paper. |

| | | | |
|---|---|---|---|
| Implementation | 10 | Who generated the random allocation sequence, who enrolled participants, and who assigned participants to interventions | See previously published Methods Paper. |
| Blinding | 11a | If done, who was blinded after assignment to interventions (for example, participants, care providers, those assessing outcomes) and how | See previously published Methods Paper. |
| | 11b | If relevant, description of the similarity of interventions | N/A. |
| Statistical methods | 12a | Statistical methods used to compare groups for primary and secondary outcomes | Methods/Results |
| | 12b | Methods for additional analyses, such as subgroup analyses and adjusted analyses | Methods/Results |
| **Results** | | | |
| Participant flow (a diagram is strongly recommended) | 13a | For each group, the numbers of participants who were randomly assigned, received intended treatment, and were analysed for the primary outcome | Event Driven Procedural Modifications |
| | 13b | For each group, losses and exclusions after randomisation, together with reasons | Event Driven Procedural Modifications |
| Recruitment | 14a | Dates defining the periods of recruitment and follow-up | Supplemental Figure: participation intervals. |
| | 14b | Why the trial ended or was stopped | All participants either withdrew, died, or completed 24 months of intervention. |
| Baseline data | 15 | A table showing baseline demographic and clinical characteristics for each group | Table 1 |
| Numbers analysed | 16 | For each group, number of participants (denominator) included in each analysis and whether the analysis was by original assigned groups | These parameters are provided for each analysis in the relevant paragraph or figure. |
| Outcomes and estimation | 17a | For each primary and secondary outcome, results for each group, and the estimated effect size and its precision (such as 95% confidence interval) | These parameters are provided for each analysis. |
| | 17b | For binary outcomes, presentation of both absolute and relative effect sizes is recommended | No binary outcomes are presented. |
| Ancillary analyses | 18 | Results of any other analyses performed, including subgroup analyses and adjusted analyses, distinguishing pre-specified from exploratory | The main statistical tests of our primary (MPI) and secondary (FAST) outcomes were pre-specified. Our other analyses can be considered exploratory. However, in the context of a full systems analysis including consideration of Hill's criteria, 'exploratory' is not necessarily pejorative, and may not substantially degrade confidence in conclusions presented. |
| Harms | 19 | All important harms or unintended effects in each group (for specific guidance see CONSORT for harms) | We identified no harms or harmful unintended effects to any participant, other than perhaps as described in the main body of the paper for the effect of the intervention on gamma-glutamyl-alpha-lysine. |

| | | | |
|---|---|---|---|
| **Discussion** | | | |
| Limitations | 20 | Trial limitations, addressing sources of potential bias, imprecision, and, if relevant, multiplicity of analyses | These are discussed in the main text. The main limitations are the low racial and ethnic diversity of the population and the small number of participants. Some analytes may have elevated significance due to regression towards the mean. |
| Generalisability | 21 | Generalisability (external validity, applicability) of the trial findings | Generalizations are discussed in the main text. Despite low racial diversity, the genetic and metabolic diversity of the population was large and the overall findings are likely to be highly generalizable. |
| Interpretation | 22 | Interpretation consistent with results, balancing benefits and harms, and considering other relevant evidence | Consistency and biological plausibility are woven into the main text. With respect to the intervention, the main side effects are beneficial (e.g., improved cardiovascular health). Harms could conceivably ensue from overconfident belief in conclusions or elements of discussion in our paper or irrational extrapolation to conclusions not present in the paper. These could lead to unnecessary expense or to opportunity cost. |
| **Other information** | | | |
| Registration | 23 | Registration number and name of trial registry | Title Page |
| Protocol | 24 | Where the full trial protocol can be accessed, if available | Attached as a Supplemental Appendix. |
| Funding | 25 | Sources of funding and other support (such as supply of drugs), role of funders | See Funding paragraph |
